# Genetic susceptibility to neurodevelopmental conditions associates with neonatal DNA methylation patterns in the general population: an individual participant data meta-analysis

**DOI:** 10.1101/2024.07.01.24309384

**Authors:** I. K. Schuurmans, D. Smajlagic, V. Baltramonaityte, A. L. K. Malmberg, A. Neumann, N. Creasey, J. F. Felix, H Tiemeier, J. B. Pingault, D. Czamara, K. Raïkkönen, C. M. Page, R. Lyle, A. Havdahl, J. Lahti, E. Walton, M. Bekkhus, C. A. M. Cecil

**Author notes:** **Correspondence:** Charlotte A.M. Cecil, Department of Child and Adolescent Psychiatry and Psychology, Erasmus MC University Medical Center Rotterdam, PO Box 2040, 3000 CA Rotterdam, the Netherlands. These authors have contributed equally (I. K. Schuurmans, D. Smajlagic).

## Abstract

**Background:** Autism spectrum disorder (ASD), attention-deficit/hyperactivity disorder (ADHD), and schizophrenia (SCZ) are highly heritable and linked to disruptions in foetal (neuro)development. While epigenetic processes are considered an important underlying pathway between genetic susceptibility and neurodevelopmental conditions, it is unclear (i) whether genetic susceptibility to these conditions is associated with epigenetic patterns, specifically DNA methylation (DNAm), already at birth; (ii) to what extent DNAm patterns are unique or shared across conditions, and (iii) whether these neonatal DNAm patterns can be leveraged to enhance genetic prediction of (neuro)developmental outcomes.

**Methods:** We conducted epigenome-wide meta-analyses of genetic susceptibility to ASD, ADHD, and schizophrenia, quantified using polygenic scores (PGSs) on cord blood DNAm, using four population-based cohorts (*n*_pooled_=5,802), all North European. Heterogeneity statistics were used to estimate overlap in DNAm patterns between PGSs. Subsequently, DNAm-based measures of PGSs were built in a target sample, and used as predictors to test incremental variance explained over PGS in 130 (neuro)developmental outcomes spanning birth to 14 years.

**Outcomes:** In probe-level analyses, SCZ-PGS associated with neonatal DNAm at 246 loci (p<9×10^−8^), predominantly in the major histocompatibility complex. Functional characterization of these DNAm loci confirmed strong genetic effects, significant blood-brain concordance and enrichment for immune-related pathways. 8 loci were identified for ASD-PGS (mapping to *FDFT1* and *MFHAS1*), and none for ADHD-PGS. Regional analyses indicated a large number of differentially methylated regions for all PGSs (SCZ-PGS: 157, ASD-PGS: 130, ADHD-PGS: 166). DNAm signals showed little overlap between PGSs. We found suggestive evidence that incorporating DNAm-based measures of genetic susceptibility at birth increases explained variance for several child cognitive and motor outcomes over and above PGS.

**Interpretation:** Genetic susceptibility for neurodevelopmental conditions, particularly schizophrenia, is detectable in cord blood DNAm at birth in a population-based sample, with largely distinct DNAm patterns between PGSs. These findings support an early-origins perspective on schizophrenia.

**Funding:** HorizonEurope; European Research Council

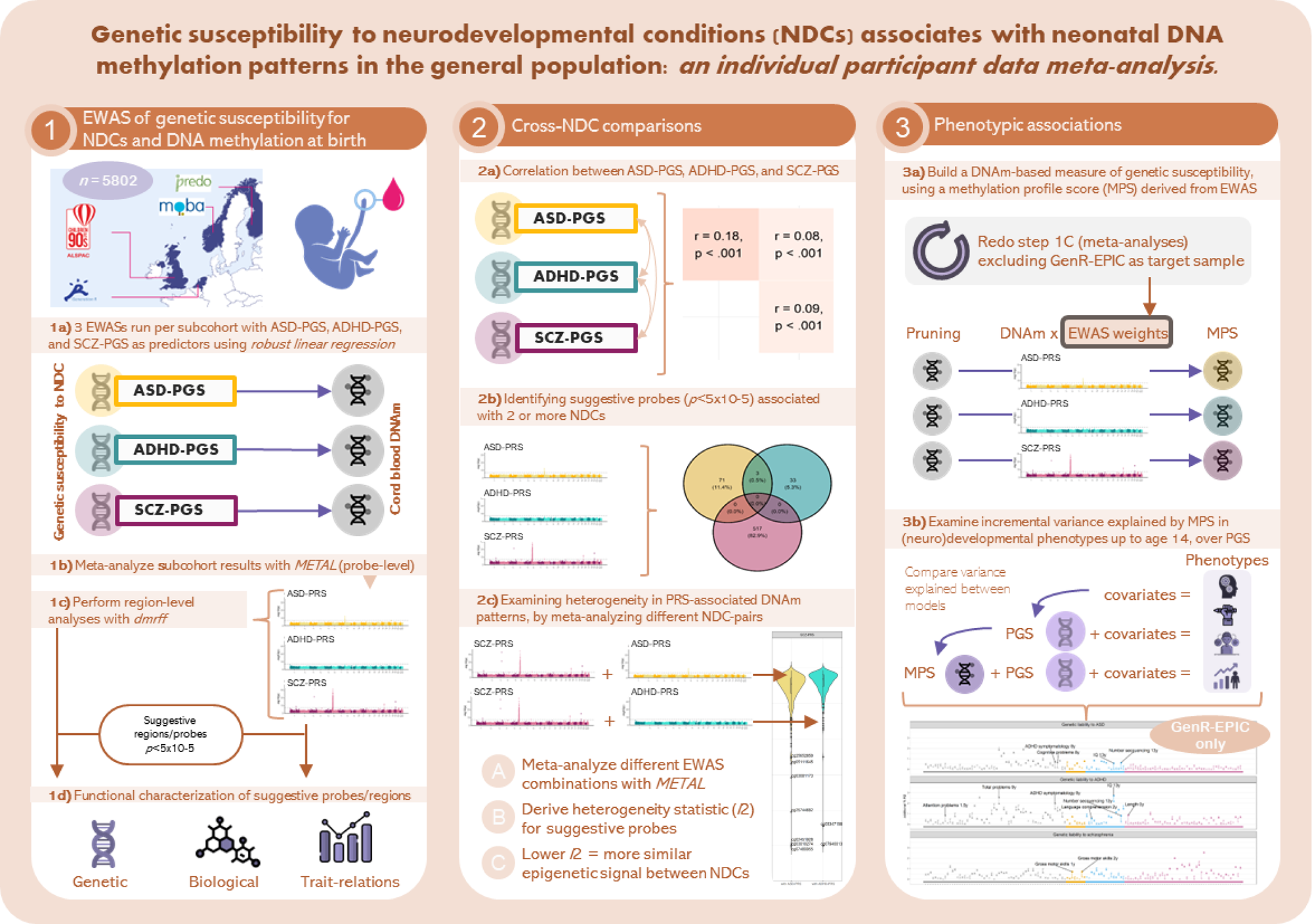

## Introduction

Neurodevelopmental conditions (NDCs) refer to a set of complex, multifactorial conditions involving perturbations in brain development that begin in foetal life.^1^ The corresponding ICD-11 diagnostic category includes in this category conditions with a developmental onset, such as autism spectrum disorder (ASD) and attention-deficit/ hyperactivity disorder (ADHD). Schizophrenia is also regarded as having neurodevelopmental origins, despite its later age of onset.^1,2^ A common feature of these three conditions is their high genetic contribution, evidenced by both family-based studies (twin-based heritability estimates ∼80%)^3–5^ as well as by large-scale genome-wide association studies (GWAS; single nucleotide polymorphism-based heritability ∼20-40%).^6^ However, the biological mechanisms underlying phenotypic presentation of these disorders remain poorly understood. Epigenetic processes that modulate gene expression, such as DNA methylation (DNAm), may be promising molecular candidates in search for biological markers and mediators of genetic and environmental influences on neurodevelopmental risk.

DNAm from peripheral blood is increasingly being used in the clinic together with genetic data for diagnosing Mendelian neurodevelopmental disorders, showing utility in differentiating complex cases with ambiguous phenotypes, compared to the use of genetic data alone.^7^ In contrast, the extent to which genetic susceptibility to *complex* (highly polygenic) NDCs associates with DNAm is less clear. So far, a limited set of studies have examined whether GWAS-derived polygenic scores (PGS) of ASD, ADHD, and schizophrenia associate with DNAm patterns, irrespective of diagnostic status (all case-control, *n*<1,300).^8–10^ Using a PGS of ASD, Hannon et al. (2018) identified genome-wide significant associations with DNAm from neonatal heel-pricks at two CpGs^8^, Mooney et al (2020) found that a PGS of ADHD was associated with saliva-derived DNAm in mid-to-late childhood at one CpG. Finally, a PGS of schizophrenia was found to associate with DNAm from adult blood at two CpGs, but those findings were not replicated in an independent cohort.^10^ For ASD and ADHD, DNAm patterns more strongly associated with PGS than with diagnostic status itself.^8,9^

While these studies provide preliminary support for a link between genetic susceptibility to NDCs and DNAm, key gaps remain. First, studies have focused exclusively on clinical case-control samples^8–10^ and associations within the *general population* have yet to be characterized. This is important given that NDCs typically manifest as a continuum, with a diagnosis of ADHD, ASD, or SCZ representing the tail end of this continuum. Furthermore, DNAm patterns are developmentally dynamic and tissue-specific,^11^ and existing studies have varied in the age and tissue of DNAm assessment (i.e., neonatal heel-pricks, saliva in childhood, blood in adulthood). Growing evidence suggests that DNAm variation at birth may be a particularly informative marker of neurodevelopmental risk, with several recent studies identifying cord blood DNAm as a stronger predictor of neurodevelopmental risk than DNAm measured in childhood. Cord blood DNAm may thus be a better proxy for congenital effects associated with NDCs, with this signal becoming noisier over time (e.g., due to postnatal exposures and immune-related changes^11^). In addition, the fact that cord blood DNAm precedes symptom onset makes it an especially promising tissue for early risk prediction, while also minimizing reverse causality. Second, studies have so far focused on PGS for individual NDCs in isolation, despite evidence of genetic and phenotypic overlap between these susceptibilities.^6^ Examining multiple PGS within the same set of individuals would offer a valuable opportunity to characterize unique versus shared epigenetic correlates of genetic risk across conditions. Finally, no research has examined the potential utility of the identified epigenetic marks in predicting (neuro)developmental outcomes. Given that PGS currently explain little variance in NDCs in the general paediatric population, examining whether incorporating additional information on genetic susceptibility from a different biological and regulatory level amplifies PGS prediction could have important implications for early risk detection.^12^

To address these gaps, we conducted a large-scale epigenome-wide association meta-analysis of genetic susceptibility to ASD (ASD-PGS), ADHD (ADHD-PGS), and schizophrenia (SCZ-PGS), leveraging individual participant data from four population-based prospective birth cohorts with DNAm obtained in the same tissue and time point (cord blood at birth) with a total combined sample size of 5,802 participants from North European datasets. Specifically, (i) we investigated epigenome-wide associations of PGSs with cord blood DNAm in the general population (probe and region level), and performed follow-up characterization to examine genetic influences (i.e., methylation quantitative trait loci (mQTL) and twin heritability estimates), associations with gene expression, blood-brain concordance, enriched functional pathways, and developmental dynamics of identified signals; (ii) we examined whether epigenetic patterns are distinct or shared across genetic susceptibilities, and (iii) we explored whether incorporating a DNAm-based measure of genetic susceptibility at birth amplifies PGS prediction of neurodevelopmental outcomes across childhood.

## Methods

### Study population

This study features four population-based birth cohorts: the Generation R Study (GenR),^13^ The Prediction and prevention of preeclampsia and intrauterine growth restriction (PREDO),^14^ the Avon Longitudinal Study of Parents and Children (ALSPAC),^15^ and the Norwegian Mother, Father, and Child Cohort Study (MoBa).^16^ Inclusion criteria and cohort-specific descriptions of methods can be found in the **Supplementary Information**. The meta-analyses included 5,802 participants of North European ancestry (**Table 1**). This study was conducted according to the Helsinki Declaration of the World Medical Association, and written informed consent was provided by all participating mothers.

**Table 1.** Population characteristics for all sub-cohorts cohorts separately.

|  | GenR<sub>450K</sub> | GenR<sub>EPIC</sub> | PREDO | ALSPAC | MoBa-1 | MoBa-2 | MoBa-4 | MoBa-8 |
| --- | --- | --- | --- | --- | --- | --- | --- | --- |
| **Cohort characteristics** |  |  |  |  |  |  |  |  |
| Cohort | GenR | GenR | PREDO | ALSPAC | MoBa | MoBa | MoBa | MoBa |
| Country | NL | NL | FI | UK | NO | NO | NO | NO |
| Sample size | 1,317 | 1,097 | 767 | 731 | 355 | 128 | 739 | 668 |
| DNAm array for cord blood | 450K | EPIC | 450K | 450K | 450K | 450K | EPIC | EPIC |
| **Child characteristics** |  |  |  |  |  |  |  |  |
| Child sex, female, n (%) | 657 (49.1) | 568 (51.8) | 362 (47.2) | 372 (50.9) | 169 (47.5) | 70 (53.9) | 362 (51.3) | 353 (54.6) |
| Gestational age at birth, weeks, mean ± SD | 40.1 ± 1.5 | 40.0 ± 1.5 | 39.8 ± 1.6 | 39.6 ± 1.5 | 39.6 ± 1.5 | 39.4 ± 1.5 | 39.6 ± 1.7 | 39.6 ± 1.9 |
| **Maternal characteristics** |  |  |  |  |  |  |  |  |
| Age, years, mean ± SD | 31.7 ± 4.2 | 31.4 ± 4.3 | 33.3 ± 5.8 | 29.8 ± 4.5 | | | | |
| Self-reported smoking during pregnancy, yes, n (%) | 275 (23.0) | 227 (22.9) | 31 (4.0) | 75 (10.3) | 40 (11.1) | 12 (10.9) | 55 (9.6) | 67 (12.2) |
| Educational level, low, n (%) | 24 (1.8) | 45 (4.2) | 335 (43.7) | 109 (15.1) | 6 (1.8) | 4 (3.3) | 12 (1.8) | 8 (1.3) |
| Educational level, medium, n (%) | 426 (32.8) | 377 (35.3) | 173 (22.6) | 457 (63.3) | 133 (33.3) | 33 (27.3) | 225 (33.4) | 199 (32.3) |
| Educational level, high, n (%) | 848 (65.3) | 645 (60.4) | 239 (31.2) | 156 (21.6) | 220 (64.9) | 84 (69.4) | 437 (64.8) | 410 (66.5) |

*Note.* NL = The Netherlands; UK = United Kingdom; FI = Finland; NO = Norway; 450K = Illumina Infinium HumanMethylation450 BeadChip; EPIC = Illumina MethylationEPIC BeadChip. There were missing values maternal characteristics variables. Self-reported smoking is shown here as more directly interpretable than prenatal maternal smoking as assessed by DNAm. Rates of low/medium/high education are not directly comparable across cohorts as educational systems differ between countries. See Supplementary Information for specific definitions.

### Genetic susceptibility for NDCs

We calculated polygenic scores with the latest GWAS summary statistics for three NDCs: ASD (ASD-PGS), ADHD (ADHD-PGS), and schizophrenia (SCZ-PGS)^17–19^, using *PRSice2* (https://choishingwan.github.io/PRSice/). First, we clumped correlated SNPs within a 250 kb window at an R^2^ threshold of 0.1. Second, PGSs were thresholded, by calculating PGSs against multiple *p*-value thresholds (only SNPs with a GWAS *p-*value below threshold are included in the PGS), and selecting the threshold for which PGS explains most variance in diagnosis-related measures across cohorts (0·5 for ASD-PGS and 0·01 for ADHD-PGS). For SCZ-PGS, a fixed threshold of 0·05 was chosen in accordance with the original GWAS,^17^ due to the lack of relevant phenotypic measures in several of the cohorts. Detailed descriptions of genotyping and PGS calculation are available in the **Supplementary Information**.

### DNA methylation

DNAm was extracted from cord blood and bisulfite-converted with the EZ-96 DNA Methylation kit (Zymo Research Corporation, Irvine, USA). Samples were run on the Illumina Infinium HumanMethylation450 BeadChip (Illumina Inc., San Diego, USA) or the Illumina MethylationEPIC BeadChip (EPIC), which include 485,577 and 867,531 CpGs, respectively. DNAm beta values were winsorised (> +/−3 SD) to reduce the influence of outliers. We excluded sites only available in one cohort and sites that are cross-reactive or polymorphic (indicated by the R package *maxprobes*; https://github.com/markgene/maxprobes), leaving 795,580 sites (380,778 EPIC-only, *n*=2,504). Cohort-specific quality control and normalization procedures are described in the **Supplementary Information**.

### Covariates

Covariates included sex, gestational age at birth, prenatal maternal smoking assessed by DNAm (for comparability across cohorts), cell-type proportions estimated via the combined cord-blood reference panel,^20^ genomic principal components to adjust for population stratification, and technical covariates to adjust for batch effects (differing per cohort; for full details, see the **Supplementary Information**).

### Analyses

**Step 1. Epigenome-wide associations.** Each cohort performed a probe-level epigenome-wide association study (EWAS) to assess associations between PGSs (ASD-PGS, ADHD-PGS, SCZ-PGS; predictors) and DNAm at birth (outcome) with covariate adjustment, separately for each NDC and CpG. We run robust linear regressions (iterated re-weighted least squares), which is less sensitive to potential heteroscedasticity and influential outliers, using the *MASS* package (version 7.3) in R version 4.1.3. Findings from individual cohorts were pooled with inverse-variance weighted fixed effects meta-analysis with *METAL* (EWAS-MA; http://www.sph.umich.edu/csg/abecasis/metal/). To assess the stability of probe-level results, a leave-one-out meta-analysis on sub-cohort was performed for the top 10 significant hits per PGS. In addition, we performed regional analyses, examining differentially methylated regions (DMRs) with the *dmrff* R package, which defines significant DMRs based on proximity, p-value, and directional consistency of CpGs (default settings). Associations were defined as genome-wide significant at a threshold of *p*<9×10^−8^ (Bonferroni-corrected for number of effective tests)^21^ and suggestive at *p*<5×10^−5^. Suggestive results were annotated using the *meffil* R package, and functionally characterized using several openly available resources (**Table 2**).

**Table 2.**
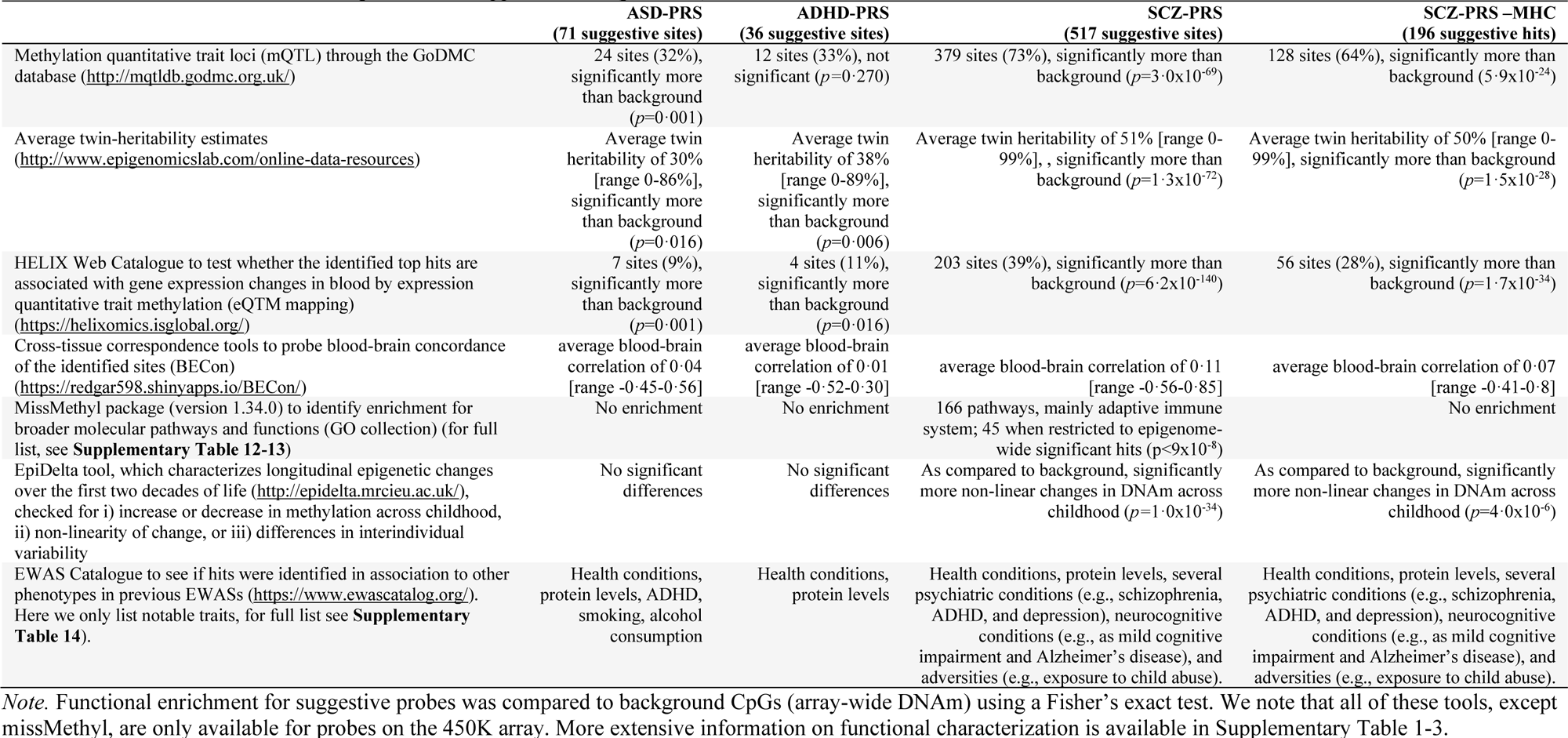
Functional characterization of probe-level suggestive sites (p<5×10^−5^).

**Step 2. Cross-NDC comparisons.** We first examined correlations between ASD-PGS, ADHD-PGS, and SCZ-PGS. Second, we identified CpGs shared across the PGS-specific EWAS-MA results, defined as CpGs showing suggestive associations with >2 PGSs. Third, we pooled together results from each EWAS-MA in pairs (e.g. EWAS-MA ASD-PGS with EWAS-MA ADHD-PGS) with inverse-variance weighted fixed effects meta-analysis (cross-NDC meta-analyses) in *METAL*. We examined heterogeneity (*I*^2^) statistics for suggestive sites, which quantifies the proportion of variance across PGS-specific EWAS-MA results attributable to heterogeneity rather than chance. Lower *I*^2^ values reflect epigenetic associations across the compared PGSs are more similar.

**Step 3. Phenotypic associations.** Finally, we built a DNAm-based measure of genetic susceptibility, by constructing methylation profile scores (MPS) for each PGS at birth (capturing the broader epigenetic signal associated with genetic susceptibility for ASD, ADHD, and schizophrenia), and tested whether these MPSs explain additional variance in (neuro)developmental outcomes over and above the PGSs themselves. To avoid over-fitting, we re-ran the EWAS-MAs removing one dataset, which we used as a target sample (GenR_EPIC_, *n*=1,097). We multiplied the EWAS-MA weights for suggestive sites (*p<*5×10^−5^) with methylation beta-values in GenR_EPIC_, performed clumping based on co-methylation patterns within GenR_EPIC_, and aggregated weighted sites into a single aggregate score similar to PGS calculation. As a baseline, we examined the incremental variance explained by PGS, above covariates, in (neuro)developmental phenotypes (spanning motor, cognitive, and behavioural domains at different time points; anthropometric variables included for comparison). Next, we evaluated incremental variance explained by MPS, above PGS and covariates. Significance was defined phenotype-wide as *p<*8×10^−4^ (Bonferroni-corrected for the number of effective tests *n*=61, Galwey method) and nominally at *p<*0·05.

Further details about step 1-3 are described in the **Supplementary Information.**

## Results

### Study characteristics

A total of 5,802 participants (50·2% female) were included in this study (**Table 1**). Gestational age at birth was similar across cohorts, however, rates of self-reported pregnancy smoking (any) ranged from 9·6% in MoBa-4 to 23·0% in GenR.

### Epigenome-wide associations of genetic susceptibility for NDCs and DNA methylation at birth

In probe-level analyses, ASD-PGS was associated with DNAm at 8 CpGs at birth after Bonferroni-correction (*p<*9×10^−8^; in total 74 suggestive at *p<*5×10^−5^), all of which were located on chromosome 8 close to the *MFHAS1* and *FDT1* genes (**Supplementary Table 1**), and nearly all present only on the EPIC array. No probe-level hits were identified for ADHD-PGS after Bonferroni-correction (36 suggestive, **Supplementary Table 2**). In contrast, the SCZ-PGS was associated with DNAm at 246 CpGs after Bonferroni-correction (517 suggestive, **Supplementary Table 3**). The majority of these sites were on chromosome 6 (96% at *p<*9×10^−8^, 87% at *p<*5×10^−5^), mostly within the major histocompatibility complex (MHC; between positions 29,640,000 and 33,120,000; 62% of Bonferroni-correction hits, 61% suggestive hits). Consistently, probe-level associations with SCZ-PGS were attenuated when the MHC was partially or fully omitted from the PGS (sensitivity analyses in GenR_EPIC_, see **Supplementary Information**). **Table 3** shows the 10 top-ranking CpGs for each PGS. EWAS-MA results showed no indication of genomic inflation (**Figure 1**), and leave-one-out results indicated that associations were unlikely driven by a single cohort (**Supplementary Figure 1**).

**Figure 1.**
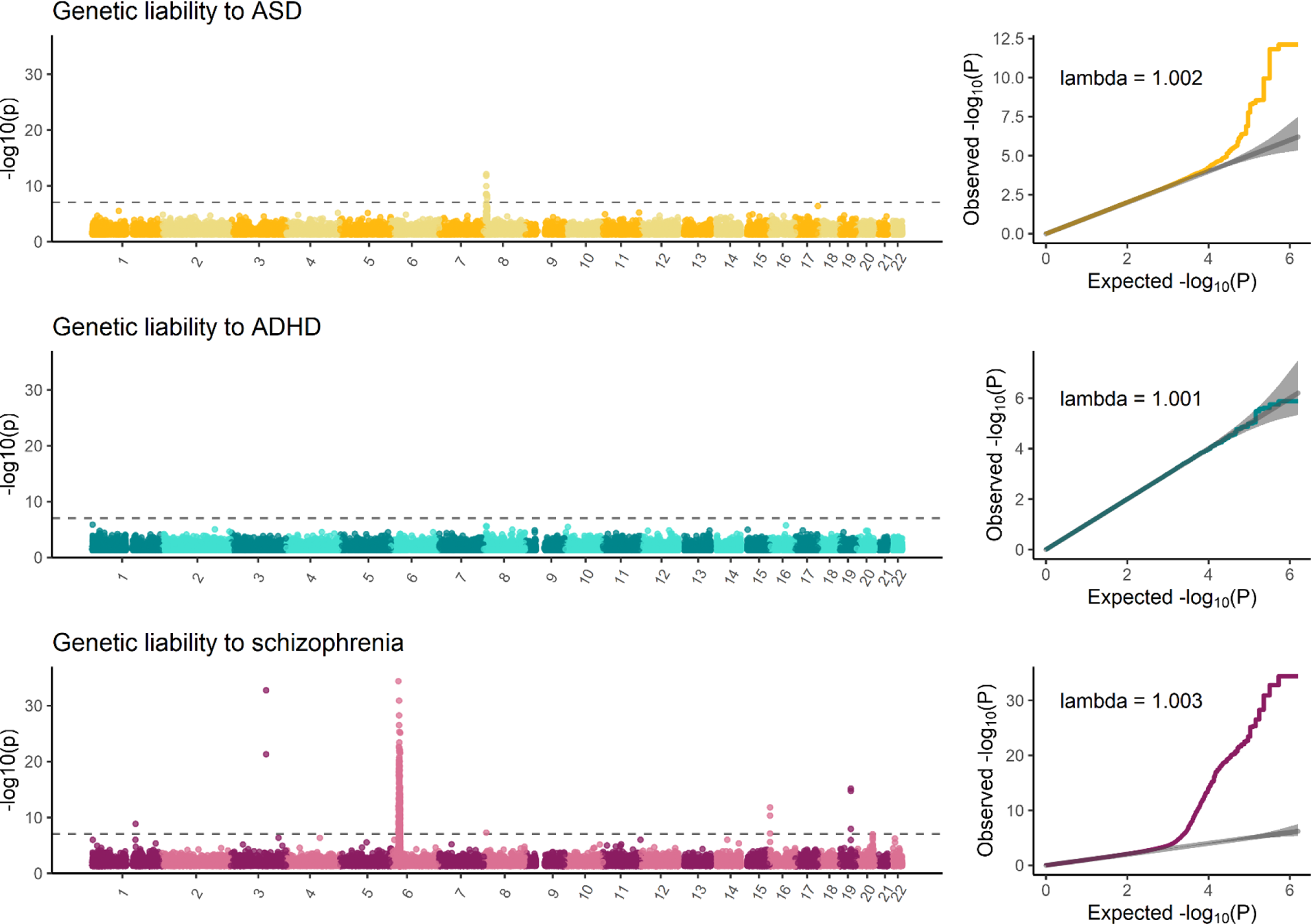
Manhattan plots and related quantile-quantile plots. Manhattan plots show which CpGs are associated with genetic susceptibility to neurodevelopmental conditions, with the grey dotted line indicating the epigenome-wide significance of p<9×10^−8^.

**Table 3.**
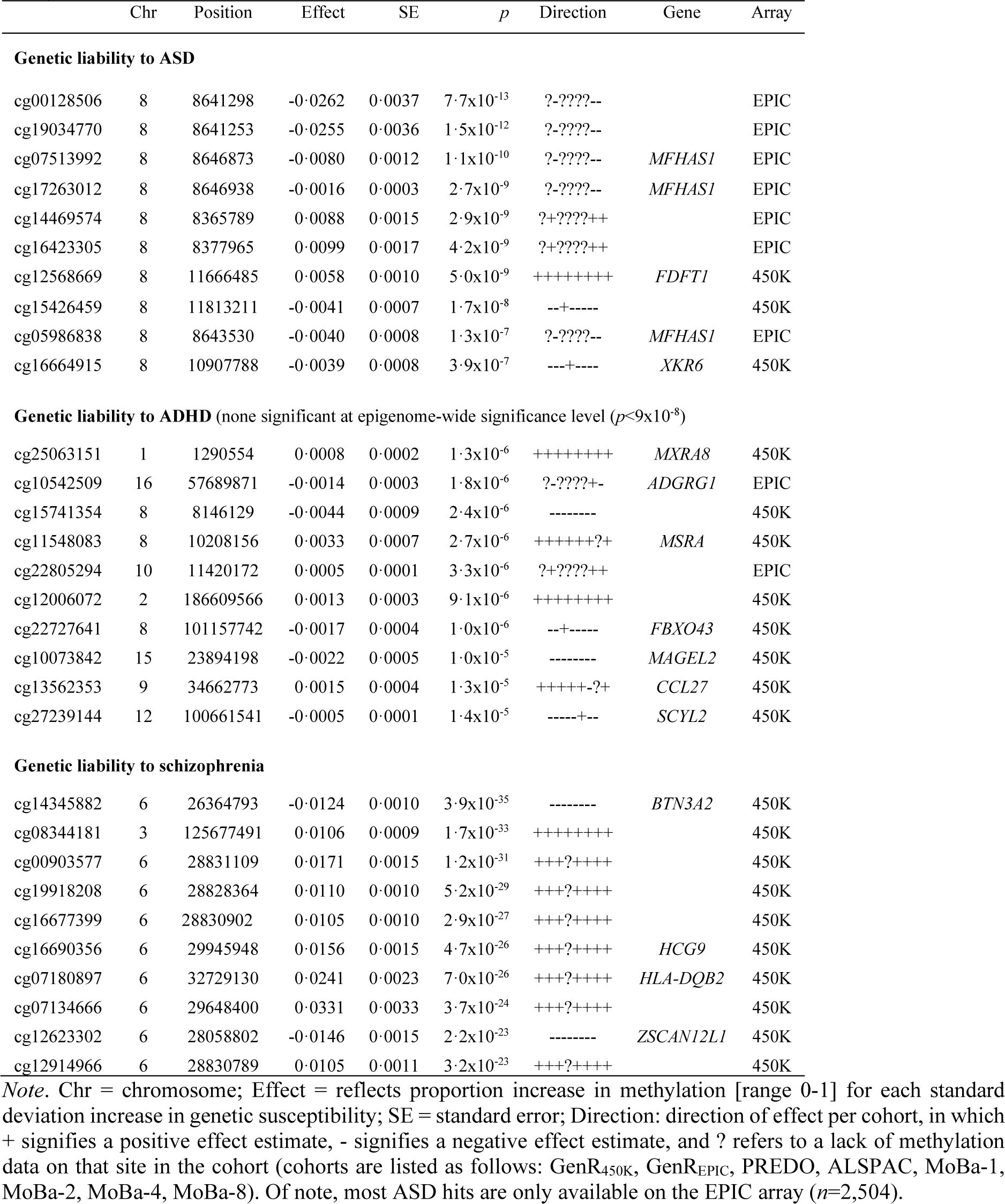
Top 10 significant loci of genetic susceptibility for neurodevelopment conditions with cord blood DNA methylation.

In region-level analyses (combining proximal CpGs into a smaller set of DNAm regions), a large number of DMRs were identified for all three PGSs, with 130 regions associated with ASD-PGS (*p<*9×10^−8^; 251 at *p<*5×10^−5^), 166 regions with ADHD-PGS (305 suggestive), and 157 regions with SCZ-PGS (297 suggestive, **Supplementary Table 4-6**). Regions overlapped only slightly with probe-level results (genes overlapping between probe-level and region-level results: ASD-PGS 5·4%; ADHD-PGS 7·4%; SCZ-PGS 12·4%).

Follow-up on probe-level and region-level analyses indicated that SCZ-PGS sites, as compared to ASD-PGS and ADHD-PGS sites, showed (i) more genetic influence (more mQTLs, higher twin heritability estimates), (ii) stronger blood-brain concordance, (iii) more enriched biological pathways (particularly related to adaptive immune response), (iv) more significant non-linear change across childhood, and (v) more often reported links to psychiatric disorders, neurocognitive conditions, and adversities, based on existing EWAS studies. These patterns were largely driven by the MHC locus because results attenuated when analyses were restricted to suggestive SCZ-PGS hits outside this locus. Full functional characterization of probe-level results can be found in **Table 2** and **Supplementary Table 1-3;** for region-level functional characterization see **Supplementary Table 7.**

### Cross-NDC comparisons

ASD-PGS and ADHD-PGS (*r*=·18) were more strongly correlated with each other than with SCZ-PGS (*r*=·08-09, **Figure 2, panel A**). Further, while probe-level methylation patterns of genetic susceptibility for NDCs were largely unrelated, three suggestive sites were shared between the ASD-PGS and ADHD-PGS (cg19034770, cg15741354, and cg11548083) but not with SCZ-PGS (**Figure 2, panel B**). A considerable proportion of variance in methylation patterns across PGSs could be attributed to heterogeneity (mean heterogeneity>74·1%; **Figure 2, panel C, Supplementary Table 8**).

**Figure 2.**
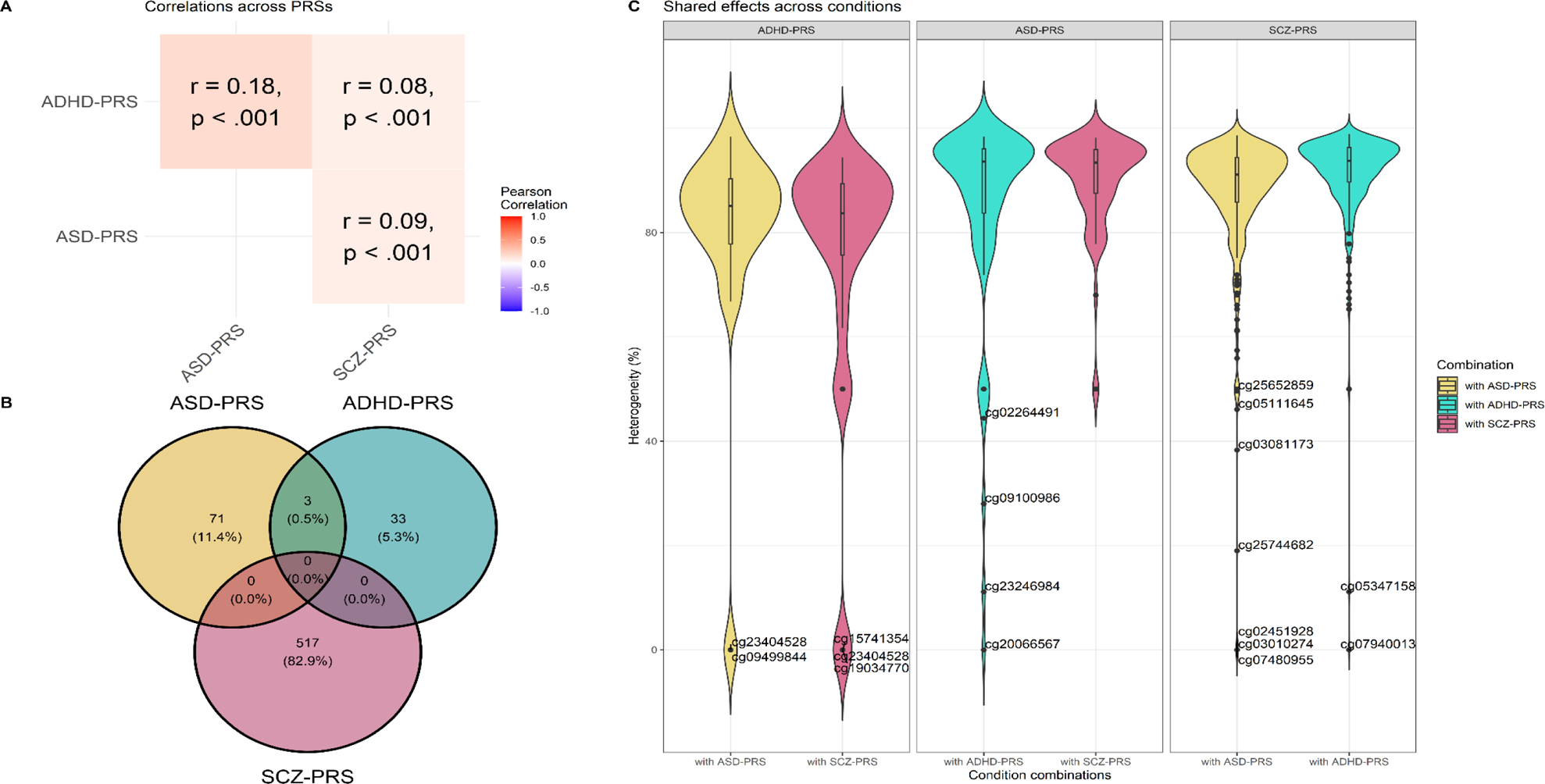
Cross-NDC comparisons for top-hits. Panel A shows correlations across genetic susceptibility (PGSs) for neurodevelopmental conditions (meta-analyzed in a fixed-effect model). Panel B shows the suggestive CpGs that are shared across conditions (p<5×10^−5^). Panel C shows heterogeneity (*I*^2^) for suggestive sites. *I*^2^ indicates the percentage of variability in associations between genetic susceptibility and DNA methylation, of a given CpGs, across two conditions, that can be attributed to variability between genetic susceptibility to the two conditions. A lower *I*^2^ value therefore reflects more similar effect sizes, for a given CpG, across the compared conditions.

### Phenotypic associations

In our target sample (GenR_EPIC,_ *n=*1,097), PGS and MPS together explained up to 2.7% of variance in phenotypes above covariates. On average, the MPS contributed to 37.2% of their total explained variance. ASD-PGS showed Bonferroni-corrected associations above covariates with 4/130 outcomes (*p<*8×10^−4^; nominally at *p<*0·05 with 20/130), ADHD-PGS with 11/130 outcomes (nominally 43/130), and SCZ-PGS with 1/130 outcomes (nominally 38/130), all above covariates. The MPS did not show any Bonferroni-corrected associations above covariates and PGS. Nominal increased variance (*p<*0·05) was found for for (i) ASD-PGS-MPS in ADHD symptoms, IQ, and number sequencing abilities, (ii) ADHD-PGS-MPS in attention and total emotional and behavioural problems, ADHD diagnosis, language comprehension, number sequencing, height, and IQ, (iii) SCZ-PGS-MPS in gross motor skills (**Figure 3**). Detailed results are shown in **Supplementary Table 9-11** and **Figure S3**.

**Figure 3.**
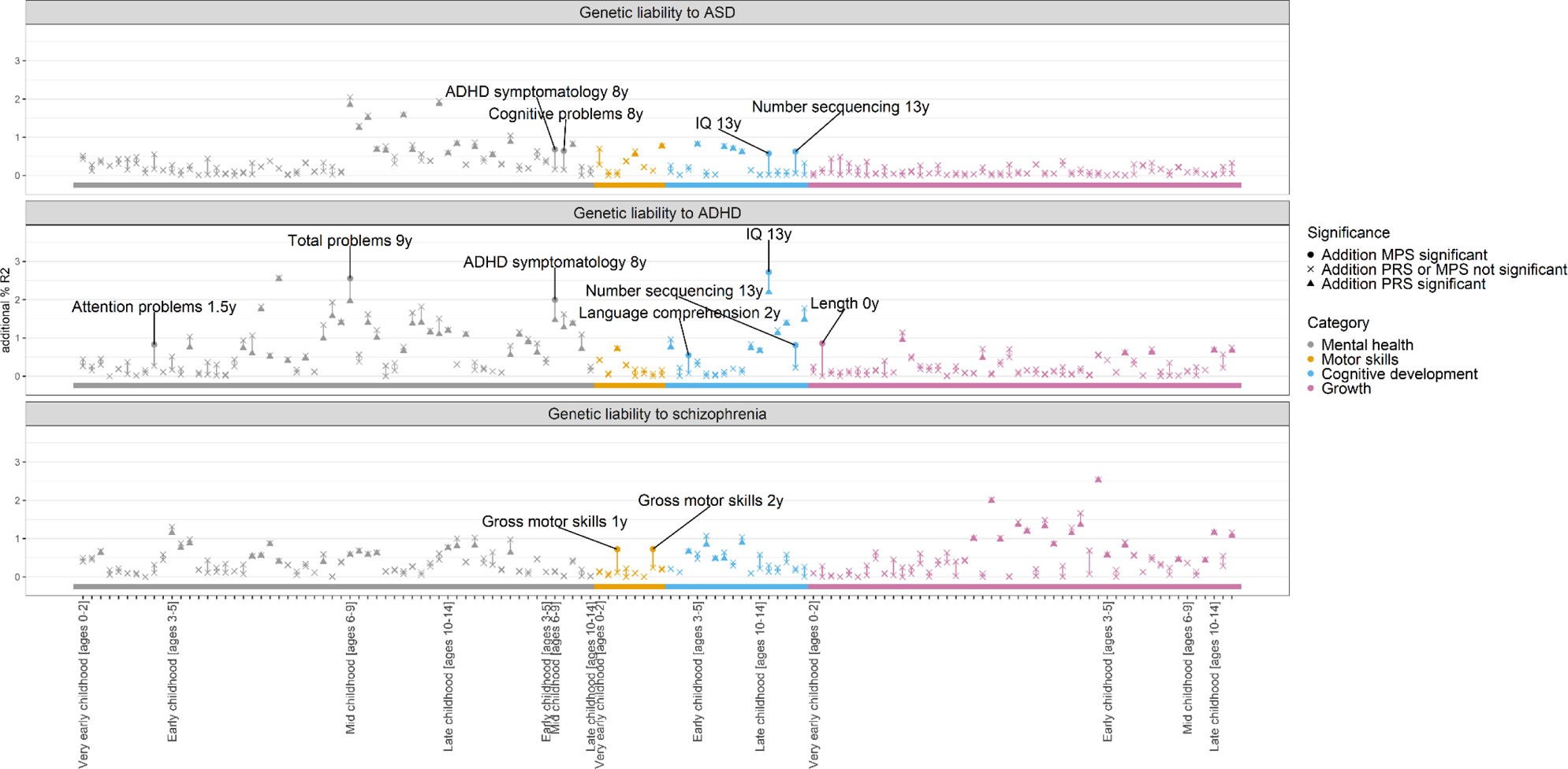
Phenotypic associations with genetic susceptibility (PGSs) to neurodevelopmental conditions and their methylation profile scores (MPS) in the target sample (GenR_EPIC_; *n=*1,097). Instances where the MPS explained additional phenotypic variance on top of the PGS (p<0·05) are indicated. More detail can be found in Supplementary Table S9-11.

## Discussion

We examined whether genetic susceptibility to NDCs is associated with DNAm patterns in cord blood, pooling data from four population-based cohorts totalling almost 6,000 participants. Our meta-analytic EWAS revealed strong probe-level associations between genetic susceptibility to schizophrenia and DNAm (246 hits), compared to ASD (8 hits) and ADHD (none). SCZ-PGS hits were mainly located within the MHC on chromosome 6, a well-established genetic risk locus for schizophrenia with a key role in immune function. In contrast to probe-level hits, region-level analyses (combining proximal DNAm sites into a smaller set of DNAm regions) detected a large number of differentially methylated regions across all three PGS (130-166 regions). PGS showed little overlap in their DNAm associations, suggesting largely distinct epigenetic signals. Finally, a DNAm-based score of genetic liability nominally increased variance in several developmental outcomes.

Our findings suggest that part of the polygenic contribution for schizophrenia is detectable epigenetically in the general population, even at a modest sample size relative to the case-control GWAS used to calculate the SCZ-PGS (*n*>200,000). Observing a strong signal already at birth (preceding by many years symptom onset) provides additional support for an early-origins perspective of schizophrenia.^2^ Previous work has found that genes implicated in schizophrenia are highly expressed in the foetal brain^17^; we extend this by showing that associations are also present early on a gene regulatory level (i.e., epigenetics). Perhaps unsurprisingly, our EWAS findings aligned closely with the discovery GWAS.^17^ The majority of identified hits clustered on chromosome 6, with 62% situated within the MHC (including our top hit cg14345882, *p*=3·9×10^−35^, annotated to the promoter region of *BTN3A2*), likely because of the strong and extended linkage disequilibrium pattern of this region. The MHC region is the strongest known genetic risk locus for schizophrenia,^22^ playing a key role in immune function (e.g., encoding proteins for antigen presentation),^23^ alongside neurodevelopmental and brain-related processes (e.g., synaptic pruning).^22^. Other EWAS signals overlapping with the schizophrenia GWAS included significant associations of SCZ-PGS with CpGs located near (i) the *ROPN1B* in chromosome 3, an important gene for male fertility and sperm motility; (ii) the *SSTR5-AS1* gene in chromosome 16, implicated in cancer development and progression;^24^ and (iii) the *RYR1* gene in chromosome 19, which encodes a ryanodine receptor found in skeletal muscle, of which mutations have emerged as a common cause of inherited neuromuscular disease.^25^ Notably, while our results reveal a widespread DNAm signal, consistent with the complex genetic architecture of schizophrenia, not all GWAS peaks were mirrored in our EWAS findings. It will be of interest in future to establish whether these may be less linked to DNAm patterns generally, whether associations may emerge later in development, or whether these are present in different tissues (e.g., brain).

Substantially fewer hits were identified in the same sample for ASD-PGS and ADHD-PGS, which may reflect limited power of the PGSs due to the smaller sample size of the discovery GWAS (ASD: 18,381 individuals diagnosed and 27,969 controls; ADHD: 38,691 individuals diagnosed and 186,843 controls; schizophrenia: 76,755 individuals diagnosed and 243,649 controls,),^17–19^ Interestingly, GWAS sample sizes do not entirely explain our pattern of findings, as we identified no hits for ADHD-PGS but 8 hits for ASD-PGS (lowest GWAS sample size). All ASD-PGS hits were located on chromosome 8 (1.0-1.9 Mb located from most prominent ASD GWAS loci *C8orf74, SOX7, PINX1*), on/ close to *FDFT1*, a gene implicated in the biosynthesis of cholesterol,^26^ and *MFHAS1*, involved in modulating the innate immune system.^27^ Unlike the stark differences in signal observed at a probe-level, we identified a similarly large number of regions (combining multiple probes) across all three PGS. This could suggest that PGS-ASD and PGS-ADHD also associate with neonatal DNAm, but that the epigenetic signal is more diffuse or that PGSs for these conditions are currently underpowered for the detection of individual CpGs. In addition, SCZ-PGS CpGs potentially were more discoverable in EWAS, as they showed higher heritability in twin-based studies, more mQTLs, and stronger associations with expression levels in blood, as compared to ASD-PGS and ADHD-PGS hits. Overall, the probe-level epigenetic signals exhibited little overlap across NDC PGSs. This may seem surprising given the known genetic (and phenotypic) correlations between ASD, ADHD, and schizophrenia. However, the magnitude of genetic correlations tends to be modest, as also observed in our data, leaving far more variance that is ‘unique’ to each condition.

We found that incorporating a DNAm-based measure of genetic susceptibility on top of PGS could considerably (yet nominally) increase explained variance in developmental outcomes. While this finding is preliminary and in need of replication, it supports continued examination into the utility of integrating information on genetic susceptibility at multiple biological layers (PGS, DNAm-based measure of genetic susceptibility) to enhance performance of early risk prediction models. It might be particularly interesting to focus on child attentional, cognitive and motor outcomes rather than emotional/behavioural symptoms or general growth parameters, as these showed largest increases in explained variance in comparison to other phenotypes. Potentially, such phenotypes are more detectable due to biological differences (i.e., a more pronounced foetal neurodevelopmental component) or measurement-related factors, for example lower measurement error and reporting bias. In addition, DNAm-based measures of genetic susceptibility to ASD and ADHD both associated prospectively with ADHD-related phenotypes, whereas the DNAm-based measures of genetic susceptibility to schizophrenia only explained additional variance in early gross motor abilities. This is consistent with previous evidence showing that PGS-SCZ correlates poorly with psychiatric symptoms in childhood,^28^ but associates with early motor abilities.^29^ Of note, our target dataset enabled us to examine phenotypes from birth to age 14 years, i.e., before typical schizophrenia onset. In future, a longer follow-up will be valuable for establishing how the neonatal DNAm-based measures of genetic susceptibility to schizophrenia may associate with relevant phenotypes (e.g. psychotic experiences) as children transition into adulthood. Importantly, only 1-10% of children will develop one of these NDCs,^30^ which raises questions about what factors interact with genetic susceptibilities to shape phenotypic expression during development, and whether DNAm may be used in this context to improve risk stratification, as it responds dynamically to both genetic and environmental influences.

Our findings need to be interpreted in light of limitations. First, potential influences of perinatal factors cannot be completely ruled out, leaving it unclear whether our findings represent direct or indirect effects of genetic susceptibility on DNAm (e.g., genetic nurture effects). Second, as we did not explicitly investigate genetic pleiotropy, observed associations may be driven by related NDCs that were not measured (e.g., intellectual disability). Third, while we consider schizophrenia as NDC, it is not included in this category within the ICD-11 and its categorization as neurodevelopmental remains a topic of debate. Fourth, our meta-analysis lacked non-European datasets limiting the extrapolation of our results across populations, and may have been influenced by selection for example the overrepresentation of healthy newborns. Finally, while we identified numerous DNAm loci associated with genetic susceptibility to NDCs, we cannot fully ascertain their functional relevance or establish their role as a potential causal pathway to NDC pathophysiology as opposed to non-causal biomarkers for genetic susceptibility.

### Conclusions

By combining genetic and epigenetic data at birth, our findings lend novel insights into the early molecular correlates of genetic susceptibility to ASD, ADHD, and schizophrenia. We identify strong associations between the PGS of schizophrenia and neonatal DNAm patterns in the general population, particularly within the MHC region, lending further support for an early-origins perspective of schizophrenia and its links to adaptive immunity. Associations of neonatal DNAm with PGS of ASD and ADHD were present but more diffuse, possibly reflecting differences in the power of the discovery GWAS. In addition, epigenetic signals at birth are largely distinct between NDC PGSs. Finally, we found preliminary evidence that the addition of methylation data may enhance genetic prediction models of neurodevelopmental outcomes.

## Data Availability

All summary data produced in the present study will be available online at FigShare upon publication in journal

## Research in context

### Evidence before this study

Neurodevelopmental conditions (NDCs) like ASD, ADHD, and schizophrenia have a strong genetic component. Epigenetic processes, such as DNA methylation, are emerging as promising molecular systems in identifying biological markers and mediators of genetic influences on these NDCs. We searched PubMed and Google Scholar for epigenome-wide association studies of PGSs of ASD, ADHD, and schizophrenia from Jan 1, 1990, to March 1, 2024. Search terms were (“methylation” OR “DNAm” OR “epigenetics”) AND (“polygenic” OR “PGS” OR “GWAS” OR “genome wide association” OR “genetic”) AND (“ASD” OR “autism” OR “ADHD” OR “attention-deficit/hyperactivity disorder” OR “schizophrenia” OR “psychosis”), restricted to articles published in English. We identified three studies that examined the link between genetic predispositions to NDCs and peripheral blood DNA methylation patterns in clinical populations (more information in **Supplement**). We did not identify any studies that were based on the general population, pooled different datasets to identify robust signals, examined cord blood at birth, compared associations between the different NDCs, or tested the potential utility of DNAm-based measures (methylation profile scores) in the prediction of (neuro)developmental outcomes.

### Added value of this study

Our epigenome-wide meta-analysis (*n*=5,802) revealed strong associations between genetic susceptibility to schizophrenia and neonatal DNAm (246 hits), compared to ASD (8 hits) and ADHD (no hits). Most schizophrenia-PGS (SCZ-PGS) hits were found in the major histocompatibility region on chromosome 6 – a well-established genetic risk locus for schizophrenia with a key role in immune function – as well as several other loci (e.g., *ROBN1B*, *SSTR5-AS, RYR1*, *SLC39A1*) consistent with the complex genetic architecture of schizophrenia. These findings indicate that genetic susceptibility for schizophrenia can in part be detected at an epigenetic level in the general population at birth (i.e., pre-symptom manifestation), in a comparatively modest sample size relative to the case-control GWAS used to calculate the SCZ-PGS (*n*>300,000). The identified epigenetic signals differed substantially between NDC PGSs, suggesting unique underlying molecular pathways. Finally, while preliminary and in need for further replication, we found that incorporating DNAm-based measures of genetic susceptibility nominally increased explained variance for selected phenotypes.

### Implications of all the available evidence

Our findings show that genetic susceptibility to NCDs associates with epigenetic patterns already at birth and within the general population. This adds to the growing body of literature supporting an early-origins perspective of schizophrenia. Further, our results suggest that incorporating information on genetic susceptibility at multiple biological layers (PGS, MPS) may help to enhance performance of early risk prediction models. Future studies are needed to establish the functional significance of identified DNAm loci, their role in NDC pathophysiology, and potential as diagnostic markers.

## Data sharing

Summary-level and analytical code data will be made available online upon acceptation of this manuscript.

## Declarations of interest

None.

## Funding

The general design of the Generation R Study is made possible by financial support from Erasmus MC, Erasmus University Rotterdam, the Netherlands Organization for Health Research and Development and the Ministry of Health, Welfare and Sport. The EWAS data were funded by a grant from the Netherlands Genomics Initiative (NGI)/Netherlands Organisation for Scientific Research (NWO) Netherlands Consortium for Healthy Aging (NCHA; project nr. 050-060-810), by funds from the Genetic Laboratory of the Department of Internal Medicine, Erasmus MC, and by a grant from the National Institute of Child and Human Development (R01HD068437). This project received funding from the European Union’s Horizon 2020 research and innovation programme (733206, LIFECYCLE; 848158, EarlyCause).

The PREDO study has received funding from the Academy of Finland, EraNet, EVO and VTR (special state subsidy for health science research), University of Helsinki Research Funds, the Signe and Ane Gyllenberg foundation, the Emil Aaltonen Foundation, the Finnish Medical Foundation, the Jane and Aatos Erkko Foundation, the Novo Nordisk Foundation, the Päivikki and Sakari Sohlberg Foundation, Sigrid Juselius Foundation, the Sir Jules Thorn Charitable Trust, and, and the HiLife Fellows Programme 2023-25.

Core support for ALSPAC was provided by the UK Medical Research Council and Wellcome (Grant ref: 217065/Z/19/Z) and the University of Bristol. GWAS data was generated by Sample Logistics and Genotyping Facilities at Wellcome Sanger Institute and LabCorp (Laboratory Corporation of America) using support from 23andMe. This publication is the work of the authors and they will serve as guarantors for the contents of this paper. A comprehensive list of grants funding is available on the ALSPAC website (). EW also received funding from the National Institute of Mental Health of the National Institutes of Health (award number R01MH113930) and from UK Research and Innovation (UKRI) under the UK government’s Horizon Europe / ERC Frontier Research Guarantee [BrainHealth, grant number EP/Y015037/1].

The Norwegian Mother, Father and Child Cohort Study is supported by the Norwegian Ministry of Health and Care Services and the Ministry of Education and Research. We thank the Norwegian Institute of Public Health (NIPH) for generating high-quality genomic data. This research is part of the HARVEST collaboration, supported by the Research Council of Norway (#229624). We also thank the NORMENT Centre for providing genotype data, funded by the Research Council of Norway (#223273), South East Norway Versjon 6.9 3 Health Authorities and Stiftelsen Kristian Gerhard Jebsen. We further thank the Center for Diabetes Research, the University of Bergen for providing genotype data and performing quality control and imputation of the data funded by the ERC AdG project SELECTionPREDISPOSED, Stiftelsen Kristian Gerhard Jebsen, Trond Mohn Foundation, the Research Council of Norway, the Novo Nordisk Foundation, the University of Bergen, and the Western Norway Health Authorities.

The work of C.A.M.C. is supported by the European Union’s Horizon 2020 Research and Innovation Programme (EarlyCause; grant agreement No 848158), the European Union’s HorizonEurope Research and Innovation Programme (FAMILY, grant agreement No 101057529; HappyMums, grant agreement No 101057390) and the European Research Council (TEMPO; grant agreement No 101039672). This research was conducted while CAMC was a Hevolution/AFAR New Investigator Awardee in Aging Biology and Geroscience Research. The work of AN is also supported by the European Union’s HorizonEurope Research and Innovation Programme (FAMILY, grant agreement No 101057529) and the European Research Council (TEMPO; grant agreement No 101039672). JBP and NC are supported by the ERC under the European Union’s Horizon 2020 research and innovation programme (iRISK; grant agreement No 863981).

The financial supporters did not influence the results of this article. The funders had no role in the study design, data collection, analysis, interpretation of the data, or writing of the report.

## Acknowledgements

The Generation R Study is conducted by Erasmus MC, University Medical Center Rotterdam in close collaboration with the School of Law and Faculty of Social Sciences of the Erasmus University Rotterdam, the Municipal Health Service Rotterdam area, Rotterdam, the Rotterdam Homecare Foundation, Rotterdam and the Stichting Trombosedienst & Artsenlaboratorium Rijnmond (STAR-MDC), Rotterdam. We gratefully acknowledge the contribution of children and parents, general practitioners, hospitals, midwives and pharmacies in Rotterdam. The generation and management of the Illumina 450K methylation array data (EWAS data) for the Generation R Study was executed by the Human Genotyping Facility of the Genetic Laboratory of the Department of Internal Medicine, Erasmus MC, the Netherlands. We thank Mr. Michael Verbiest, Ms. Mila Jhamai, Ms. Sarah Higgins, Mr. Marijn Verkerk and Dr. Lisette Stolk for their help in creating the EWAS database. We thank Dr. A. Teumer for his work on the quality control and normalization scripts. We also thank all the research nurses, research assistants, and laboratory personnel involved in the PREDO study. We are also extremely grateful to all the families who took part in the ALSPAC study, as well as for the midwives for their help in recruiting them, and the whole ALSPAC team, which includes interviewers, computer and laboratory technicians, clerical workers, research scientists, volunteers, managers, receptionists and nurses. We thank all the children and their parents for participation. We are grateful to all the participating families in Norway who take part in this on-going cohort study, MoBa. Finally, we would like to thank prof. dr. Roel Ophoff for his helpful input on the study.

## Notes

### Competing Interest Statement

The authors have declared no competing interest.

### Author Declarations

The Medical Ethical Committee of Erasmus MC, University Medical Center Rotterdam gave ethical approval for this work Ethics Committee of Obstetrics and Gynecology at Hospital District of Helsinki and Uusimaa gave ethical approval for this work ALSPAC Ethics and Law Committee and the Local Research Ethics Committees gave ethical approval for this work the Regional Committee for Ethics in Medical Research, the Norwegian Data Inspectorate and the Institutional Review Board of the National Institute of Environmental Health Sciences, USA gave ethical approval for this work

