## Supplement for "Genetic susceptibility to neurodevelopmental conditions associates with neonatal DNA methylation patterns in the general population: an individual participant data meta-analysis"

### Introduction

Epigenetic processes that modulate gene expression, such as DNA methylation (DNAm), may be promising molecular candidates for biological markers and mediators of genetic and environmental influences on neurodevelopmental risk. However, only very few studies have examined whether GWAS-derived polygenic scores (PGS) of ASD, ADHD, and schizophrenia associate with DNAm patterns, irrespective of diagnostic status.[7-9] Using a PGS of ASD, Hannon et al. (2018) identified genome-wide significant associations with DNAm from neonatal heel-pricks at two CpG sites (cg02771117 and cg27411982) in 629 individuals diagnosed with ASD and 634 controls.[7] Mooney et al (2020) found that a PGS of ADHD associated with DNAm from saliva in mid-to-late childhood at one site (cg15472673), at genome-wide significance in a sample of 302 participants diagnosed with ADHD and 170 controls.[8] Finally, a PGS of schizophrenia associated with DNAm from adult blood at two CpGs (cg05110828 and cg03879918) in a study with 32 participants diagnosed with schizophrenia and 307 controls, although the findings were not replicated in an independent cohort (413 individuals diagnosed with schizophrenia, 430 controls).[9] In the case of ASD and ADHD, DNAm patterns were found to be more strongly associated with PGS than diagnostic status itself.[7,8]

### Methods

#### Study population

Across all studies, we included newborns if they had genetic information and information on DNA methylation (DNAm) from cord blood. In case of siblings, we included only 1 child per mother (the sibling with most data was kept, or if equal, one sibling was kept at random).

#### The Generation R Study

The Generation R Study (GenR) is a population-based prospective cohort from fetal life onwards.[1] In short, pregnant women were eligible for study participation if they were residents of the study area (Rotterdam, the Netherlands), and if they had a delivery date between April 2002 and January 2006. A total of 9,778 mothers were enrolled. These mothers, their children and partners took part in several research waves. GenR was approved by the Medical Ethical Committee of Erasmus MC, University Medical Center Rotterdam.

#### Prediction and Prevention of Preeclampsia and Intrauterine Growth Restriction

The Prediction and Prevention of Preeclampsia and Intrauterine Growth Restriction (PREDO) study is a prospective, multicenter study of Finnish women who were pregnant between 2005 and 2009 and their children.[2] PREDO recruited 1,079 women with a singleton, intrauterine pregnancy, who visited antenatal clinics at any of the 10 study hospitals for their first routine ultrasound screening at 12 to 13 weeks of gestation, of whom 969 had one or more and 110 had none of the known risk factors for preeclampsia and intrauterine growth restriction. *PREDO* was approved by the Ethics Committee of Obstetrics and Gynecology at Hospital District of Helsinki and Uusimaa.

#### The Avon Longitudinal Study of Parents and Children

The Avon Longitudinal Study of Parents and Children (ALSPAC) study is a prospective birth cohort, which invited pregnant women resident in Avon, UK with expected dates of delivery between 1st April 1991 and 31st December 1992. [3,4] The initial number of pregnancies enrolled was 14,541· Please note that the study website contains details of all the data that is available through a fully searchable data dictionary and variable search tool (http://www.bristol.ac.uk/alspac/researchers/our-data/). Participants for the current study were derived from the Accessible Resource for Integrated Epigenomic Studies (ARIES).[5] A sub-set of 1018 mother-offspring pairs were included in ARIES and were selected based on availability of DNA samples at two time points for the mother (at an antenatal clinic and at a follow-up clinic when their offspring were mean age 15·5 years) and three time points for the offspring [at birth, childhood, (mean age 7·5 years) and adolescence at mean age 15·5 years]. Ethical approval for the study was obtained from the ALSPAC Ethics and Law Committee and the Local Research Ethics Committees. Consent for biological samples has been collected in accordance with the Human Tissue Act (2004).

#### The Norwegian Mother, Father, and Child Cohort Study

The present study will use data from the Norwegian Mother and Child Cohort Study (MoBa), population-based pregnancy cohort study conducted by the Norwegian Institute of Public Health.[6,8] Participants were recruited from all over Norway from 1999 to 2008· The women consented to participation in 41% of the pregnancies. The cohort now includes 114,500 children, 95,200 mothers and 75,200 fathers. Blood samples were obtained from both parents during pregnancy and from mothers and children (umbilical cord) at birth. MoBa is also linked to the Medical Birth Registry (MBRN), a national health registry containing information about all births in Norway. The current study is based on version 12 of the quality-assured data files released for research in January 2019· The establishment of MoBa and initial data collection was based on a license from the Norwegian Data protection agency and approval from the Regional Committees for Medical and Health Research Ethics. The MoBa cohort follows regulations according to the Norwegian Health Registry Act. All MoBa participants have already provided their informed consent and have been approved by REK (REK- 2009/1899-7) and the Norwegian Data Inspectorate. Sensitive data will be protected by state-of-the art system for storage of sensitive data at Oslo University (TSD). The current study was approved by The Regional Committees for Medical and Health Research Ethics (REK-185800). Finally, *MoBa* was approved by the Regional Committee for Ethics in Medical Research, the Norwegian Data Inspectorate and the Institutional Review Board of the National Institute of Environmental Health Sciences, USA.

#### Genetic susceptibility for neurodevelopmental conditions

#### Cohort-specific genotyping

##### *The Generation R Study*

Children's blood samples were collected from the umbilical cord at birth, or by venipuncture at around age 6 if cord blood was not available. Samples were preserved at −80 °C (umbilical) or −20 °C (venipuncture) and DNA was extracted using the Qiagen FlexiGene Kit. Genotyping was conducted on Illumina HumanHap 610 or 660 Quad chips with raw data processed by Illumina Genome Studio software, applying a no-call threshold of 0·15 and a quality metric exclusion threshold of 97·5%. We merged data from both HumanHap chip types using SNPs common to both arrays, followed by rigorous quality control (QC) in PLINK. Marker QC filters were applied for call rate (<0·2 initially, <0·05 in a second, more stringent round), minor allele frequency (MAF≤0·001), differential missingness ($p$<10x10$^{-7}$), and Hardy-Weinberg equilibrium ($p$<10x10$^{-7}$). Sample QC checks included duplicate detection, sex discordance (using chromosome x heterozygosity), genotype call rate (with 95% initial threshold, 97·5% after marker QC), and high heterozygosity rate, set at over 4 SD from the mean. Imputation utilized the 1000 Genomes phase 3 data, adhering to parameters from the phasing procedure in build 37, and included both autosomal and x chromosome markers, with separate imputations for males and females. A final round of QC was performed when converting variant call format files to bed files, as we additionally excluded multi-allelic SNPs, duplicate variants, structural variants (inserts and deletions), low imputation quality ($R^2$ < 80%) and minor allele frequency (MAF < 1%).

#### *Prediction and Prevention of Preeclampsia and Intrauterine Growth Restriction*

Fetal cord blood samples were collected according to standard procedures. DNA was extracted at the National Institute for Health and Welfare, Helsinki, Finland, and the Finnish Institute of Molecular Medicine, University of Helsinki, Finland. Genotyping was performed on Illumina Human Omni Express Exome Arrays containing 964,193 SNPs. Only markers with a callrate of at least 98%, a MAF of 1% and a p-value for deviation from HWE ($p$<10x10$^{-6}$) were kept in the analysis. Samples with a callrate below 98% (n=11) were removed. Any sample-pair with IBD estimates > 0·125 was checked for relatedness. For most pairs, high IBD estimates could be explained due to African origin of these participants. As we will correct for admixture in our analyses, these participants were kept except for one pair which could not be resolved, from this pair, one participants was excluded from further analysis. Participants showing discrepancies between phenotypic and genotypic sex (n=1) were removed. We also checked for heterozygosity outliers but found none. Before imputation, AT and CG SNPs were removed. Imputation was performed using shapeit2 and impute2. Chromosomal and base pair positions were updated to the 1000 Genomes Phase 3 reference set, allele strands were flipped where necessary. After imputation, we re-ran QC, filtering out SNPs with an info score<0·8, a minor allele frequency below 1% and a deviation from HWE with a $p$<10x10$^{-6·}$ After that, data was converted to best guessed genotypes (setting the probability to at least 90%) and QC'ed again.

#### *The Avon Longitudinal Study of Parents and Children*

Genetic data were derived using the Illumina HumanHap550 quad chip from cord blood. Genotype calls were made using Illumina GenomeStudio software. QC filters were applied for a MAF cut-off at 0·01, HWE-value threshold at $p$<10x10$^{-6}$, SNP missingness at a rate of 0·01 (i.e., removing SNPs with a high proportion of missing data), individual missingness at 0·02, and finally by removing indels. The dataset was carefully checked for duplicate variants, which could arise during genotyping or imputation processes, to ensure each SNP was represented uniquely. Extreme heterozygosity rates had previously been removed by the ALSPAC team. Genetic data was imputed against the 1000 Genomes Phase 1 version 3 reference panel using Impute V2·2·2·

#### *The Norwegian Mother, Father, and Child Cohort Study*

Genotyping was performed in the following 3 projects: (i) HARVEST (~33,000 individuals genotyped on Illumina HumanCoreExome array), (ii) ERC HARVEST (~27,000 individuals Illumina Global Screening Array) and (iii) NORMENT (> 100,000 individuals genotyped on Illumina OmniExpress array). Genotyping and imputation are described in detail elsewhere (https://www.biorxiv.org/content/10·1101/2022·06·23·496289v2). Overall, MoBa consists of 26 genotyping batches grouped by 3 genotyping arrays prior imputation.

Since the imputation was done in different time points, we used ~98,00 out of ~200,000 MoBa individuals that were available to us during the duration of this project. In total, we used 8 genotyping batches (harvest12, harvest24, Norment_Jan2015, Norment_Jun2015, Norment_May2016, Norment_Feb2018, ROTTERDAM1, and ROTTERDAM2) grouped by 3 genotyping arrays named HCE (referring to Illumina HumanCoreExome array), GSA (referring to Illumina Global Screening Array), and OMNI (referring to Illumina OmniExpress array). An additional post-imputation QC was performed on the HCE, GSA, and OMNI batches. In summary, QC filters were applied for SNPs (call rate 0·02; MAF 0·01; difference in the MAFs between genotyping batches $p<10\text{x}10^{-3}$; HWE $p<10\text{x}10^{-6}$; SNPs with >0·01 Mendelian errors were removed), and individuals (call rate 0·02; inbreeding coefficient ≥0·20; unrelated individuals were ensured to have *PI_HAT* measure <0·2 by removing one individual from each pair of unrelated individuals; families with >0·05 Mendelian errors were removed). Parents and children that were imputed in different batches were combined together and QC-ed as one batch named CROSS. This resulted in 4 MoBa batches for which the polygenic scores were calculated individually and adjusted for the first 10 principal components and sex. The residuals were then combined together and used instead of polygenic scores in further analyses.

### Polygenic risk score calculation process

Genetic single nucleotide polymorphism (SNP) data were used to generate polgenic scores for three neurodevelopmental conditions: autism spectrum disorder (ASD; ASD-PGS), attention-deficit/hyperactivity disorder (ADHD; ADHD-PGS), and schizophrenia (SCZ-PGS). SNPs were identified in the most recent GWASs.[10-12] The GWAS for ASD used a sample of 18,381 cases and 27,969 controls, which identified 3 genome-wide significant SNPs.[12] The GWAS for ADHD investigated a larger sample size of 38,691 individuals with ADHD and 186,843 controls, and identified 27 genome-wide significant SNPs.[13] The largest sample size was incorporated in the schizophrenia GWAS, with a cohort of 76,755 schizophrenic individuals and 243,649 controls [10]. This study identified 313 independent genome-wide significant SNPs.

### Parameter optimization

The ASD-PGS, ADHD-PGS, and SCZ-PGS were computed using a clumping and thresholding approach in PRSice2, in order to ensure comparability of PGSs across cohorts.[14] We employed default options, meaning correlated SNPs were clumped within a 250 kb window at a $R^2$ threshold of 0·1. The ASD-PGS and ADHD-PGS were calculated at the following *p*-value thresholds: 1, 0·5, 0·4, 0·3, 0·2, 0·1, 0·05, 0·01, 0·001, $3·5\text{x}10^{-5}$, $1\text{x}10^{-5}$, $1\text{x}10^{-6}$, $1\text{x}10^{-7}$, $5\text{x}10^{-8}$, and $1\text{x}10^{-8}$. To determine the optimal threshold for PGS scores (i.e., the threshold at which the score was most strongly associated with a diagnoses-related measure), we examined the associations between PGSs at each threshold and their diagnoses-related measure. The diagnoses-related measures for ASD were assessed in GenR with an 18-item abridged Social Responsiveness Scale at age 6,[15-17] in ALSPAC using a binary question "Have you ever been told that your child has autism, Asperger's syndrome or autistic spectrum disorder?" at age 9, in PREDO using the Autism Spectrum Screening Questionnaire at age 5, and in MoBa with the Childhood Asperger Syndrome Test (CAST) at age 5.[18] The ADHD diagnoses-related measures were assessed in GenR with the Child Behavior Checklist (CBCL/6-18) inattention syndrome scale at age 10,[19] in ALSPAC at with DSM-IV diagnoses of ADHD based on the Development and Well-Being Assessment (DAWBA) questionnaires at age 10,[20] and in PREDO and MoBa with the Conners Rating Scale at age 5.[21] The diagnoses-related measure for schizophrenia was only available for ALSPAC but not for other cohorts due to the young ages examined. Instead of performing parameter optimization for this PGS, we therefore applied a pre-selected threshold of 0·05, in accordance with prior research, which performed leave-out-prediction on 98 cohorts, finding that the median p-value threshold that maximized the out-of-sample prediction in the left-out cohort was 0·05.[10]

Associations between PGSs at all threshold and diagnosis-related measures were then pooled across cohorts and meta-analyzed by implementing an inverse-variance weighted fixed effects meta-analysis using METAL to select the optimal threshold for the ASD-PGS and ADHD-PGS [22].The ASD-PGS was most strongly associated with ASD at a *p*-value threshold of 0·5 (meta-analysis of association between PGS and phenotype $p$=1·2x$10^{-6}$) for the ADHD-PGS this was at a *p*-value threshold of 0·01 (meta-analysis between PGS and phenotype $p$=9·8x$10^{-34}$ **(Supplementary Table 15).**

### DNA methylation

Each cohort calculated the methylation betas, representing the ratio of methylated signal relative to the sum of methylated and unmethylated signal measured per CpG, and normalized these betas. DNAm betas were winsorized (> +/- 3 SD) to reduce the influence of potential outliers. Cohort-specific QC and normalization procedures are described below.

### Cohort-specific DNAm processing

#### *The Generation R Study*

DNAm was collected from cord blood directly after birth. DNA was extracted using the salting-out method and was then bisulfite-converted using the EZ-96 DNAm Kit (Shallow) from Zymo Research Corporation. GenR has two batches of epigenetic data that were treated separately in analyses: (1) the Illumina Infinium HumanMethylation450 BeadChip ($GenR_{450K}$) and (2) the MethylationEPIC v1·0 BeadChip ($GenR_{EPIC}$). These batches are referred to as GenR-450K and GenR-EPIC, respectively. Both arrays are from Illumina Inc., San Diego, USA. In short, each cohort calculated the methylation betas, representing the ratio of methylated signal relative to the sum of methylated and unmethylated signal measured per CpG, and quantile normalized these betas. The data underwent QC procedures as prescribed by the CPACOR workflow, utilizing R software for data processing. All arrays were scrutinized for call rate, with those having a rate above 95% for the 450k array and 96% for the EPIC array being taken forward. Finally, we excluded samples with technical failures, such as failed bisulfite conversion, hybridization, or extension problems, as well as sex mismatch determined by the xand Y chromosome probe intensities.

#### *Prediction and Prevention of Preeclampsia and Intrauterine Growth Restriction*

DNAm was collected from cord blood directly after birth. DNAm was assessed using the Illumina Infinium HumanMethylation450 BeadChip. Samples were randomized across 96-well plates considering gender and maternal preeclampsia risk factors. Following bisulfite conversion using the EZ-96 DNAm Kit, methylation levels were reported as beta values. The QC pipeline was set up using the R-package *minfi*.[23] Three IDs were excluded as they were outliers in the median intensities. Furthermore, 20 IDs showed disconcordance between phenotypic sex and estimated sex and were excluded. Nine IDs were contaminated with maternal DNA and were also removed.[24] Methylation beta-values were normalized using the funnorm function. We excluded any probes on chromosomexor Y, probes containing SNPs and cross-hybridizing probes according to Chen et al.[25] and Price et al. [26]. Furthermore, any 5'-C-phosphate-G—3' CpGs with a detection *p*-value > 0·01 in at least 25% of the samples were excluded. After normalization two batches, i.e., slide and well, were significantly associated and were removed iteratively using the ComBat method.

#### *The Avon Longitudinal Study of Parents and Children (ALSPAC)*

DNAm was collected from cord blood directly after birth. DNAm was assessed using the Illumina Infinium HumanMethylation450 BeadChip post-bisulfite conversion with the EZ-96 DNAm Kit (shallow). The QC process was conducted using the *meffil* package in R version 3·4·3· QC steps included checks for genotype and sex mismatches, incorrect relatedness, sample concordance at different time points, dye bias, and probe detection efficiency [27].

#### *The Norwegian Mother, Father, and Child Cohort Study*

DNAm was measured in cord blood after birth in 4 different MoBa epigenetic batches (more about met001, met002, met004, and met008 is written here: https://github.com/folkehelseinstituttet/mobagen/wiki/Methylation). First two batches (met001 and met002) had the DNAm assessed using the Illumina Infinium HumanMethylation450 BeadChip, while the second two batches (met004 and met008) had the DNAm assessed using the Illumina Infinium MethylationEPIC BeadChip. The QC was performed in each batch separately, using the *minfi* package in R software. The detailed description of each QC step can be found here: https://github.com/folkehelseinstituttet/mobagen/wiki/MethylationQC. In this project, we treated these 4 batches separately and for the clarity, in the manuscript we named them MoBa-1, MoBa-2, MoBa-4, and MoBa-8.

#### Covariates

As covariates, we included child sex, gestational age at birth, prenatal maternal smoking as a score based on methylation profiles (to ensure comparability across cohorts), cell-type proportions estimated via the combined cord-blood reference panel,[28] genomic principal components to adjust for population stratification, and technical covariates to adjust for batch effects.

#### Prenatal maternal smoking as a score based on methylation profiles

To ensure comparability across cohorts, we measured maternal smoking with a DNAm maternal smoking score. This approach was based on previous work, and combines probes related to maternal smoking as found in previous epigenome-wide studies.[29-31]

#### Cell-type proportions

Cell-type proportions in cord blood methylation were estimated using the method described by Gervin et al. [28], identifying CD8T, CD4T, NK, Bcell, Mono, Gran, and nRBC.

### Other cohort-specific covariates

#### *The Generation R Study*

Gestational age at birth was ascertained through detailed ultrasound examinations performed during prenatal visits. The sex of the child was determined at birth by attending midwives. Cell-type proportions in cord blood methylation were estimated using the method described by Gervin et al.,[28] identifying CD8T, CD4T, NK, Bcell, Mono, Gran, and nRBC. The analysis accounted for population stratification by incorporating the first five genetic principal components derived from genotyped data.[32] Batch effects were addressed by including the sample plate as a covariate.

#### *Prediction and Prevention of Preeclampsia and Intrauterine Growth Restriction*

Gestational age at birth and child's sex was derived from the Finnish Medical Birth Register provided data. Cell-type proportions in cord blood methylation were estimated using the method described by Gervin et al. [28], identifying CD8T, CD4T, NK, Bcell, Mono, Gran, and nRBC. To correct for population stratification, the first three principal components were corrected for. These were derived from genotyped data. Batch effects were corrected using the ComBat method from the *sva* package in R.

#### *The Avon Longitudinal Study of Parents and Children (ALSPAC)*

Gestational age and sex were established at birth by midwives. The analysis accounted for population stratification by incorporating genetic principal components derived from genotyped data; we corrected for the first five genetic principal components. To correct for batch effects, ten surrogate variables were calculated using the *meffil* package [27]; some were subsequently removed for their association with the outcomes of interest. Specifically, eight surrogate variables were used for ADHD and nine for ASD and SCZ.

#### *The Norwegian Mother, Father, and Child Cohort Study*

Sex and gestational age were obtained from Medical Birth Registry of Norway. To correct for population stratification, ten genetic principal components were used. To correct for batch effects, surrogate variables were calculated using the *meffil* package [27]; the ones associated with the outcomes of interest were removed.

### Descriptive variables

As descriptive variables, we included maternal age at birth, self-reported maternal smoking (as self-reported smoking is shown here as more directly interpretable than DNAm scores of prenatal smoking exposure), and maternal education. These three were all cohort-specific.

### Cohort-specific descriptive variables

#### *The Generation R Study*

Maternal age was collected from mothers at the time of their inclusion in the study through self-reporting [1]. Self-reported smoking status (as in Table 1) was reported by mothers during pregnancy, and categorized into two groups: (1) those who smoked during pregnancy and (2) those who ceased smoking upon discovering their pregnancy. Lastly. Lastly, maternal education was assessed by a questionnaire and defined by the highest attained educational level and classified into three categories: low (no education or primary education), middle, (intermediate general school, vocational training) and high (Bachelor's degree, higher academic education, PhD).

#### *Prediction and Prevention of Preeclampsia and Intrauterine Growth Restriction*

Maternal age was obtained from the Finnish Medical Birth Register. Maternal self-reported smoking status (as in Table 1) was based on data recorded in the Finnish Medical Birth Register and originally classified into none, vs quit during the first trimester, vs continued smoking after the first trimester. For the current study, smoking status was dichotomized into never smoked vs. ever smoked. Self-reported level of education was measured during pregnancy, classified into primary, secondary, lower tertiary, or upper tertiary: for the current study, categories primary and secondary were combined due to a relatively small number of individuals belonging to primary category, resulting in three categories: low (primary or secondary) vs. medium (lower tertiary) vs. high (upper tertiary).

#### *The Avon Longitudinal Study of Parents and Children (ALSPAC)*

Maternal age at birth was self-reported. The maternal self-reported smoking status (as in Table 1) was captured with a dichotomous variable indicating the presence (1 or more cigarette) or absence of smoking (0 cigarettes) during pregnancy. Maternal education levels were categorized into three strata, reflecting the low (secondary education, vocational), medium (O level or A level), and high (university degree).

#### *The Norwegian Mother, Father, and Child Cohort Study*

Maternal age at birth and smoking during pregnancy were obtained from Medical Birth Registry of Norway. Maternal age at birth was recorded according to the following categories: 17 or younger, 18-19, 20-24, 25-29, 30-34, 35-39, 40-44, or 45 or older. Mothers' self-reported smoking during pregnancy was deducted from two measurements: smoking during pregnancy and smoking at the end of pregnancy. Both measurements were categorized into "no smoking", "sometimes", and "daily". If the value of smoking during pregnancy or smoking at the end of pregnancy was "sometimes" or "daily", we recoded mothers' self-reported smoking during pregnancy as "yes"; otherwise, we recoded it as "no". Information about maternal education was collected using self-reported questionnaires at 15th week of gestation. The highest level of completed education was classified according to the following categories: 9-year secondary school, 1-2 years of high school, 3-year high school, bachelor's degree, or university degree of 4 years or more.

### Analyses

#### Step 1· Epigenome-wide associations

#### *Differential methylated regions*

We examined differentially methylated regions (DMRs) using the *dmrff* package. Significant differentially methylated regions were defined based on the following criteria: (1) two neighboring CpGs can be at most 500 base pairs apart from each other; (2) CpGs have nominal EWAS $p$ values < 0·05, and (3) CpGs are in the same direction.  We employed default settings and epigenome-wide correction thresholds as per the package. To annotate and functionally characterize these regions, we first identified the CpGs comprising regions based on chromosome, start, and end positions, and then proceeded to characterize this specific set of CpGs functionally.

#### *Functional characterization*

Functional characterization was performed using a range of openly accessible resources. We performed three main types of functional characterization: (i) genetic, (ii) functional, and (iii) developmental. For genetic characterization we identified mQTLs (i.e., the GoDMC database; http://mqtldb.godmc.org.uk/);[33] and we investigated twin-heritability estimates (https://www.epigenomicslab.com/online-data-resources/).[34] For biological characterization, we included the HELIX Web Catalogue to test whether the identified top hits are associated with gene expression changes in blood by expression quantitative trait methylation mapping (eQTM; https://helixomics.isglobal.org/).[35] We also used cross-tissue correspondence tools to probe blood-brain concordance of the identified sites (BECon; https://redgar598·shinyapps.io/BECon/).[36] In addition, we implemented the missMethyl package (version 1·34·0[37]) to identify enrichment for broader molecular pathways and functions (GO collection). For characterization of developmental dynamics, we performed a look up of our suggestive hits in the EpiDelta tool, which characterizes longitudinal epigenetic changes over the first two decades of life (http://epidelta.mrcieu.ac.uk/). [38] Functional enrichment for suggestive probes was compared to background DNAm sites (array-wide DNAm, 450K only) using a Fisher's exact test. We note that all of these tools, except missMethyl, are only available for probes that are present on the 450K array.

### Step 2· Cross-condition comparisons

#### *Cross-condition meta-analyses for examining heterogeneity*

To examine whether  epigenetic signals are unique vs shared between the PGSs for ASD, ADHD and SCZ, we meta-analyzed all possible pairs of PGS-specific EWAS MA results into cross-condition meta-analyses (i.e., ASD with ADHD, ASD with schizophrenia, ADHD with schizophrenia), to capture heterogeneity across PGS-specific methylation patterns. In traditional meta-analyses, heterogeneity statistics are used to capture cross-*cohort* heterogeneity. Here, we use heterogeneity statistics to gauge cross-*condition* heterogeneity. This method mirrors the strategy of Gidziela et al. (2023), who used heterogeneity analysis to explore etiological proximity between various neuropsychiatric conditions at a genetic level.[39] We estimated the extent of heterogeneity using the $I^2$ statistics, which quantifies the proportion of variation across PGS-specific EWAS-MA results attributable to heterogeneity rather than random chance.[40] As such, this measure reflects whether confidence intervals for a given CpG is independent across PGS-specific EWAS-MAs. Using heterogeneity analysis has advantages overusing pairwise correlation tests of meta-analyzed weights, as it avoids biases introduced by (1) the predominance of PGS-SCZ hits, as it might skew results due to the increased number of data points, and (2) the exclusive consideration of effect sizes, which might overlook the variability of those effect sizes (e.g., some non-significant probes may have large effect sizes, as well as large standard errors, which is not reflected in the magnitude of effect size only). We interpret heterogeneity for all suggestive sites at $p$<5x10$^{5}$ (e.g., interpreting heterogeneity for all suggestive sites with ASD-PGS; evaluating whether these sites share signal with PGS-ADHD and PGS-SCZ).

### Step 3· Phenotypic associations

#### *Methylation profile scores*

We constructed methylation profile scores (MPSs) of genetic susceptibility to ASD, ADHD and SCZ in order to assess whether they can enhance PGS prediction of neurodevelopmental phenotypes. This follow-up analysis was performed specifically in the $GenR_{EPIC}$ cohort as the target sample, as it is the largest participating cohort with DNAm data analyzed on the EPIC array (enabling thus to use in the construction of MPSs CpG sites from both the 450k and EPIC arrays). MPSs were calculated using the most comparable approach to the one used for our PGS calculation; that is, by summing the weighted DNAm values of selected CpGs, where the weights are the effect sizes from the EWAS. We specifically employed the P+T CoMeBack approach, using (1) independent EWAS-level summary statistics, (2) co-methylation pruning, and (3) p-value thresholding.[41] As such, first, we derived summary statistics from an *independent* EWAS (i.e., not including the target data, here $GenR_{EPIC}$) to prevent overfitting; we therefore re-meta-analyzed summary statistics for each PGS EWAS without the $GenR_{EPIC}$ dataset. Second, we *pruned* the DNAm data, using CoMeBack,[42] which involves linking adjacent array probes in the DNAm data of the testing dataset or a reference panel based on specific criteria: (1) two probes are less than 2kb apart; (2) the reference human genome annotation shows a set of unmeasured genomic CpGs between them; (3) the density of unmeasured genomic CpGs between them is at least one CpG every 400bp. A comethylated region is identified if all pairs of adjacent probes exhibit a correlation square ($R^2$) greater than 0·3. Only the CpG site with the most significant *p*-value within each comethylated region is retained. Third, the EWAS summary statistics were *p-value-thresholded* (i.e., only including CpG sites that reached an EWAS-level *p*-value below those thresholds into your MPS). Often, MPSs are calculated against several different p-value thresholds, which are then evaluated in a separate independent dataset to determine which threshold explains most variance in the phenotype. However, evaluating different thresholds in an independent dataset requires additional sample size, which means we lose power. We therefore instead used a predetermined threshold at $5x10^{-5}$ (reflecting our suggestive threshold), including 58 CpGs for the ASD-PGS, 37 for the ADHD-PGS, and 530 for the SCZ-PGS. After pruning, the ASD-PGS-MPS included 51 CpGs, ADHD-PGS-MPS included 32 CpGs, and the SCZ-MPS included 272 CpGs. Weights for all three MPSs are available in **Supplementary Table 16-18.** Of note, MPSs were significantly correlated with PGSs for all three conditions in $GenR_{EPIC}$(PGS-ASD: $r$=0·17, $p$<0·001; PGS-ADHD: $r$=0·14, $p$<0·001; PGS-SCZ: $r$=0·23, $p$<0·001).

#### *Phenotypes*

We tested associations between MPSs and 130 outcomes. These spanned (neuro)developmental phenotypes, including motor, cognitive, and behavioral outcomes, as well as anthropometric outcomes for comparative purposes (given that anthropometry captures development, but does not tap specifically into a neurodevelopment). All phenotypes were measured between birth up to age 14, and were often repeated across time (e.g. attention problems at 1·5, 3, 6, 10, and 14 years). For all measures, we included validated subscales and total scales, as described in detail below.

#### Motor outcomes

Child Development Inventory. The Child Development Inventory (CDI) was used to assess language and motor skills (CDI) at the aged 6 months, 1 year, 1·5 years, and 2 years.[43] We measured 4 different subscales: the expressive language scale (15 yes/no items), the language comprehension scale (15 yeas/no items), the fine motor development scale (30 yes/no items), and the gross motor development scale (30 yes/no items). Age-appropriate items were selected (i.e., items indicating development of a child up to 2 years old). The score for the scale can be calculated as the total number of items marked yes for the scale.

#### Cognitive outcomes

Brief Rating Inventory of Executive Function-Preschool Version. The Brief Rating Inventory of Executive Function-Preschool Version (BRIEF-P)[44] was administered at age 4 to measure functions related to inhibition, shifting, emotional control, working memory and planning in preschool children in everyday life. The BRIEF-P consists of 63 items forming five subscales: inhibition (16 items), shifting (10 items), emotional control (10 items), working memory (17 items), and plan/organizing (10 items). Main caregivers indicated the extent to which their child had displayed the behaviour during the last month on a 3-point Likert scale (1=never, 2=sometimes, 3=often). Scale scores reflect the mean scores on the relevant items, with higher scores indicating more problems in executive functions.

Snijders-Oomen Niet-verbale intelligentie test. We used two subtests of the validated Dutch non-verbal intelligence test: Snijders-Oomen Niet-verbale intelligentie test 2·5-7 jaar revisie (SON-R 2·5-7) to measure nonverbal intelligence[45] at age 5. The subtests were Mosaics, which evaluates spatial insight, and Categories, which examines abstract reasoning abilities. Raw scores were transformed into nonverbal IQ scores, with the use of age-specific normal values based on the child's age.

Cito. School performance was assessed with the Dutch standardized end-of-primary-school test, created by the Central Institute for Test Development (CITO; www.cito.com) at age 12. The test assesses different areas, including language, mathematics, and world orientation (optional), and gives an indication of the child's intelligence, motivation, concentration, and drive to learn. It is administered in the final year of primary school in class, just before children move on to secondary school, usually around age 11-12 years. Test scores were obtained from CITO data linkage and, if not available, retrieved from maternal reports by questionnaire.[46]

Wechsler Intelligence Scale for Children - Fifth Edition. The Wechsler Intelligence Scale for Children - Fifth Edition (WISC-V) was used to assess IQ[47] at age 13· A subset of tests was administered, namely the Matrix reasoning to measure fluid reasoning, Digit Span for working memory, Coding for processing speed, and Vocabulary for verbal comprehension. Raw scores were standardized to t-scores using age- and sex specific norm groups. Then, the sum of the t-scores were converted to IQ, using a conversion table that has 0·93 correlation with full-scale IQ.[48]

#### Behavioral outcomes

Child Behavior Checklist. The Child behavior Checklist (CBCL) was used to measure emotional and behavioral problems during the past six months at the ages 1·5 years, 3 years, 5 years, 9 years, and 14 years. Age-appropriate versions of the instruments were used: the preschool version (CBCL/1·5-5)[49] to the age of 6 and the school-age version (CBCL/6-18)[19] for the ages after that. The instrument has more than 100 items (100 for the preschool age version, 113 for the school-age version), using a 3-point Likert scale (0=not True, 1=somewhat/sometimes true, 2=often/very true). Items can be grouped into two main categories, internalizing problems and externalizing problems. We additionally included the empirically derived subscales of the CBCL, also known as the Syndrome Scales, which encompass behaviors characteristic of internalizing and externalizing conditions. The computation for total scales and subscales differs over version.

Social Responsiveness Scale. Parents completed the Social Responsiveness Scale (SRS) to measure ASD-like behaviors for children covering the past six months[15-17] at age 5· A total of 18 individual questions were scored on a 4-point Likert scale (0=not true, 1=sometimes true, 2=often true, 3=almost always true. DSM-5 symptom domains for ASD included in the SRS are social communication/interaction and restricted/repetitive patterns of behaviour, interests, or activities.

Conners Scale. The Conners' Parent Rating Scale–Revised Short Form (Conners, Sitarenios, Parker, & Epstein, 1998) was used to assess attention problems in children[21] at age 8. This questionnaire contains 27-items that were rated on a 4-point Likert scale (1=not true at all, never, seldom, 2=just a little true, occasionally, 3=pretty much true, often, quite a bit, 4=very much true, very often, very frequent). This version of the Conners' Parent Rating Scale includes 4 scales: Cognitive Problems/Inattention (6 items), Hyperactivity (6 items), Oppositional (6 items), and the ADHD index (ADHDi; 12 items).

Psychotic like experiences. Lifetime experience of delusions was self-reported at 14 years old, using 6 items relating specifically to delusional experiences drawn from the Kiddie-Schedule for Affective Disorders and Schizophrenia (K-SADS; Kaufman et al., 1997; Townsend et al., 2020) which was adapted for use in a self-reported questionnaire.[50] Items were rated on a 3-point Likert scale from (1=not true, 2=yes, likely, 3=yes, definitely).

#### Anthropometrics outcomes

For child anthropometrics, we measured height, weight, and BMI (weigh/heigh2), as captured during research visits at the Generation R center at birth, 2 months (0·1 years), 3 months (0·25 years), 4 months (0·33 years), 8 months (0·7 years), 1 year, 1·2 years, 1·8 years, 2·1 years, 2·5 years, 2·8 years, 3·3 years, 4·2 years, 5 years, 9 years, 13 years.

#### *Analytical strategy*

Using linear regression analyses, we first defined our baseline, for which we examined the incremental variance explained by PGS, over and above covariates, for each neurodevelopmental and anthropometric phenotype. Next, we evaluated the incremental variance explained by MPS, over and above PGS and covariates. Models were compared using analysis of variance. To ensure multiple testing was taken into account, we applied the Bonferroni correction, specifically adjusting for the effective number of independent tests. We employed the Galwey method[51] to estimate the number of independent test ($n_{eff}$=61), as this method is often applied when outcomes are dependent, for example due to correlations between phenotypes or the use of repeated measures for a given phenotype. The method estimates the number of effective tests based on the eigenvalues of the correlation matrix across outcomes, using the following formula:

$$m = \frac{(\sum_{i=1}^{k} \sqrt{\lambda_i'})^2}{\sum_{i=1}^{k} \lambda_i'}$$

Consequently, the adjusted p-value threshold for significance was set as 0·05/61=8x10$^{-4}$.

### Results

#### Epigenome-wide associations between genetic susceptibility for neurodevelopmental conditions and DNA methylation at birth

#### Polygenic risk scores for schizophrenia, omitting the MHC

To further explore the role of the MHC region, we revisited the SCZ-PGS EWAS, introducing *two* new SCZ-PGSs that omit SNPs within the MHC locus (chr6:25-35k). For the first PGS (PGS-SCZ$_{Rank1}$), we excluded the MHC region (6:25,000,000-35,000,000) while preserving the rank 1 region (6:28,303,247-28,712,247; reflecting the broader surrounding area of rs115329265). For the second PGS (PGS-SCZ$_{Variant}$), we excluded the MHC region (6:25,000,000-35,000,000) while preserving on variant 6:28,712,247 (rs115329265). Here, we followed the exclusion approach as outlined by the GWAS from the PGC-schizophrenia working group in 2023.[10] This approach is more stringent as compared to PGS-SCZ$_{Rank1}$.

Next, we reran the EWAS  using either  the PGS-SCZ$_{Rank1}$ and PGS-SCZ$_{Variant}$ (rather than the PGS-SCZ as applied in the main analyses) within GenR$_{EPIC}$ as a sensitivity analysis. For PGS-SCZ, we identified 6 EWAS-level hits ($p$<9x10$^{-8}$; 151 suggestive at $p$<5x10$^{-5}$). Of those, 2 were located in the MHC ($p$<9x10$^{-8}$; 53 suggestive at $p$<5x10$^{-5}$). For both PGS-SCZ $_{Rank1}$ and PGS-SCZ$_{Variant}$, we found 10 EWAS-level hits ($p$<9x10$^{-8}$; 168 suggestive at $p$<5x10$^{-5}$). None of those were within the MHC ($p$<9x10$^{-8}$; 10 and 1 suggestive at $p$<5x10$^{-5}$, respectively). As expected, we observed an attenuation of results in the MHC region for both PGS-SCZ$_{Rank1}$ and PGS-SCZ$_{Variant}$, as also evident in **Supplementary Figure 2.**

**Supplementary Figure 1.** Forest plot showing associations between PGS of each neurodevelopmental conditions and DNA methylation for the 10 most significant associations. Note that for each CpG, the top line (pink triangle), represents the meta-analyzed result. The rows below show results when leaving out one cohort at a time (circle, other colors). Cohorts are presented in order of sample size (large at the top).

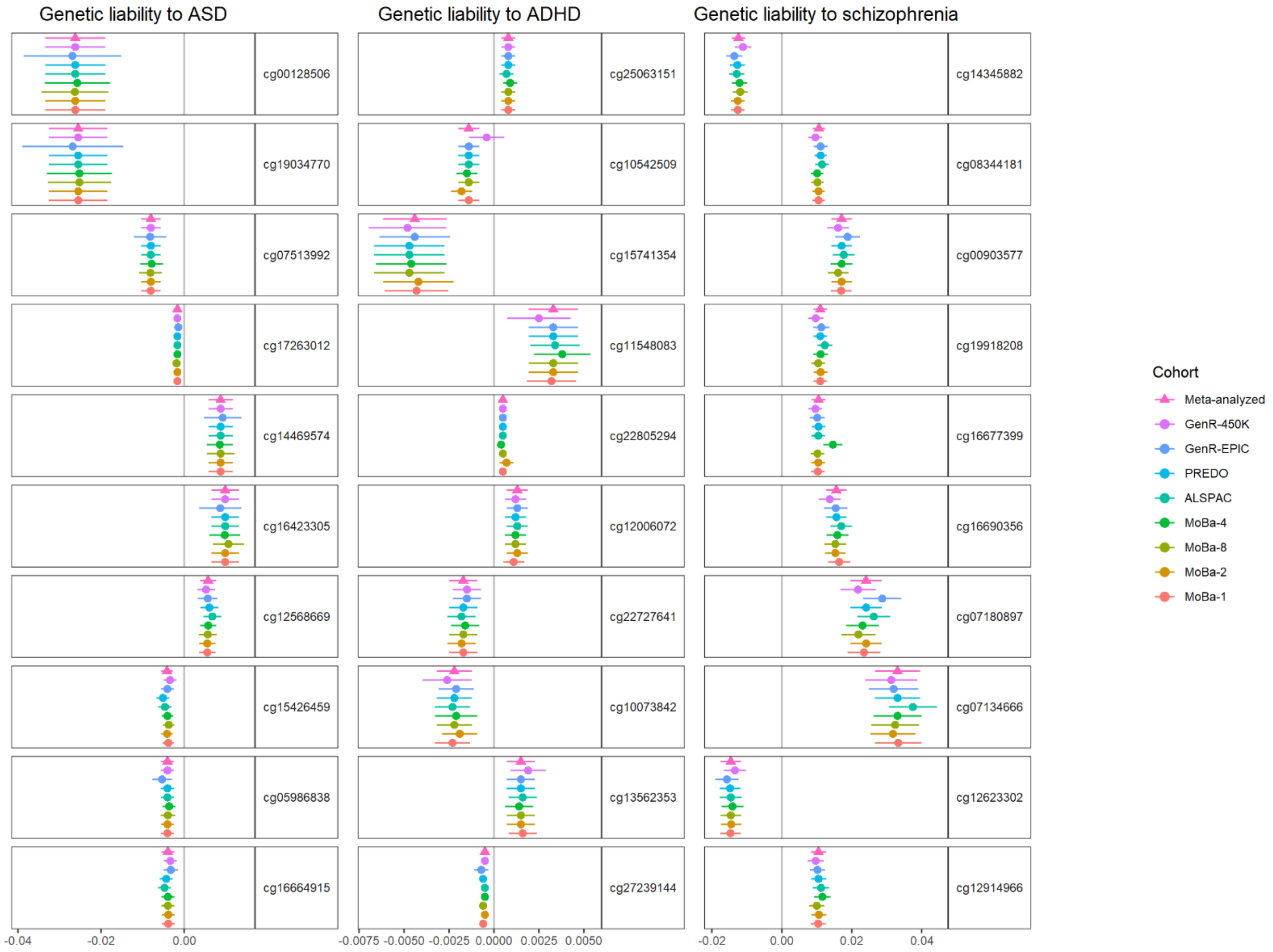

**Supplementary Figure 2.** Manhattan plots show associations between cord blood DNAm and genetic susceptibility to schizophrenia when using different approaches to account for the MHC region in the SCZ-PGS. Results are from sensitivity analyses conducted $GenR_{EPIC}$ only. The grey dotted line indicates the epigenome-wide significance threshold of $p<9x10^{-8}$. Upper panel present results for SCZ-PGS, calculated using only clumping (as applied in main analyses). Middle panel presents results for $PGS\text{-}SCZ_{Rank1}$, where we excluded the MHC region (6:25,000,000-35,000,000) while preserving the rank 1 region (6:28,303,247-28,712,247; reflecting the broader surrounding area of rs115329265). Bottom panel presents results for $PGS\text{-}SCZ_{Variant}$, where we excluded the MHC region (6:25,000,000-35,000,000) while preserving on variant 6:28,712,247 (rs115329265).

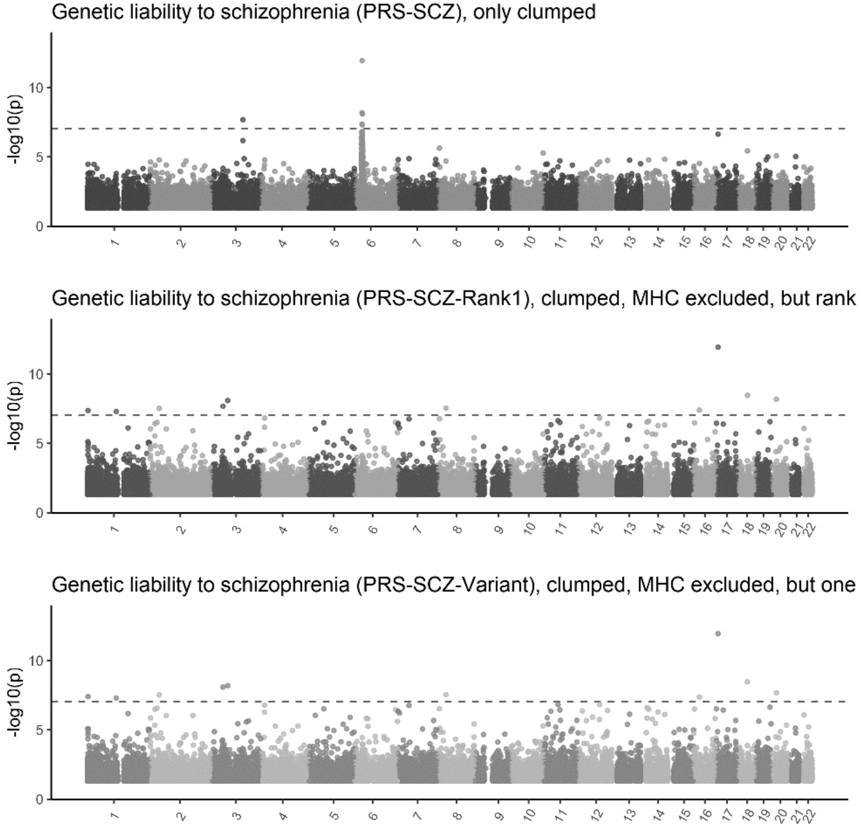

**Supplementary Figure 3.** Phenotypic associations with genetic susceptibility (PGS)to neurodevelopmental conditions and their methylation profile scores (MPS) in the target sample ($GenR_{EPIC}$; *n*=1,097). Instances where the MPS explained additional phenotypic variance on top of the PGS (p<0·05) are indicated.

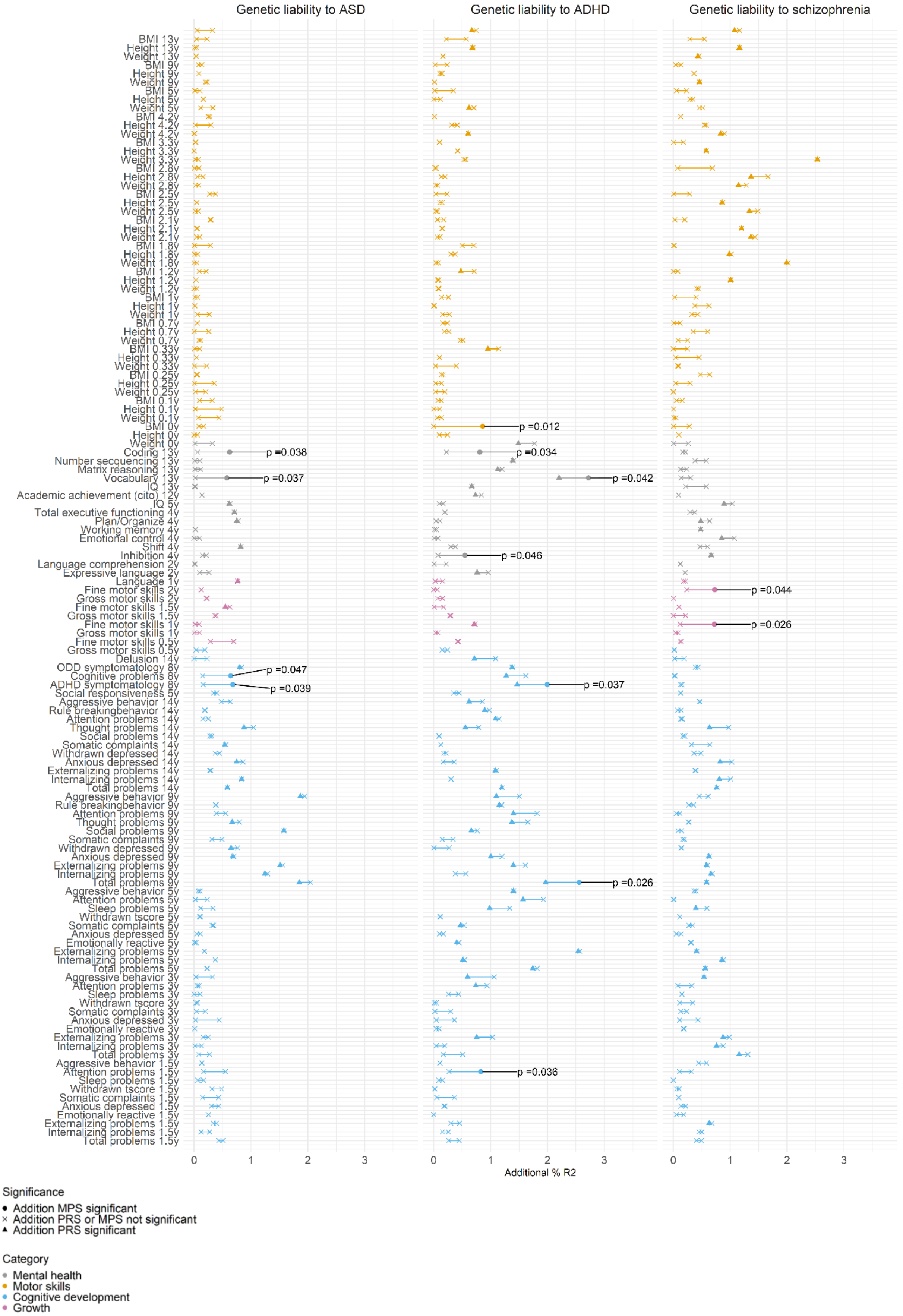
